# Programmable Fluorescent Aptamer-Based RNA Switches for Rapid Identification of Point Mutations

**DOI:** 10.1101/2025.03.07.25323576

**Authors:** Zhaoqing Yan, Yudan Li, Amit Eshed, Kaiyue Wu, Zachary M. Ticktin, Vel Murugan, Efrem S. Lim, Fan Hong, Alexander A. Green

## Abstract

The ability to detect single-nucleotide polymorphisms (SNPs) is critical for identifying genetic disorders, assessing pathogen drug resistance, and preventing infection transmission. Achieving a delicate balance across sequence-specific recognition, RNA structural stability, and functional efficacy based on SNP-induced changes in RNA structure is crucial to precise genotyping using RNA-based probes. Here, we report an in silico-designed aptamer-based RNA switch we term FARSIGHT (for **<u>F</u>**ast **<u>A</u>**ptamer-based **<u>R</u>**eporter for **<u>SI</u>**ngle-nucleotide-specific **<u>G</u>**enotypying through **<u>H</u>**ybridiza**<u>T</u>**ion) that provides rapid, low-leakage, and multiplexed identification of virtually any target sequence in as little as 5 minutes with single-nucleotide specificity. Coupling FARSIGHTs with nucleic acid sequence-based amplification (NASBA) enables robust detection of single-nucleotide mutations at attomolar concentrations with strong fluorescence output. To evaluate these assays, we deploy them to distinguish the SARS-CoV-2 Omicron variant from other SARS-CoV-2 variants (Alpha, Beta, and Gamma) with 100% accuracy on RNA extracted from clinical saliva samples, as confirmed by reverse transcription quantitative polymerase chain reaction and genomic sequencing. FARSIGHTs can thus be easily reprogrammed for genotyping new pathogens with pandemic potential, with potential uses in point-of-care settings for monitoring of emerging infectious diseases and for personalized healthcare applications.

## INTRODUCTION

Single-nucleotide polymorphisms (SNPs) and single-nucleotide variants (SNVs) are crucial factors influencing a variety of biological processes ranging from protein function^1^ to disease susceptibility^2^, response to medication^3^, and evolutionary adaptation^4^. Mutations occur frequently in the genome and have far-reaching effects on cell function. Meanwhile, small mutations can drive virus adaptation through genetic changes that abolish the target sites of antiviral drugs and improve viral fitness. Indeed, new variants of severe acute respiratory syndrome coronavirus 2 (SARS-CoV-2) have been reported to be 70% more contagious than earlier ones^5,6^ and studying the emergence and propagation of these mutations can be helpful in understanding evolution and disease mechanisms. Despite the profound impact of single-nucleotide changes, detecting these minute changes can be challenging without sophisticated instrumentation.

Genomic sequencing and microarray technologies have been widely used for detecting genetic mutations ^7–9^. Yet, these methods are typically limited to central laboratories because of the need for complex equipment, expensive reagents, and trained personnel. Clustered regularly interspaced short palindromic repeats (CRISPR)-based diagnostics hold a promising for detecting genetic variations^10^, but they require that mutations are located near a PAM region ^11–13^ or within a narrow window^14–17^ in the targeting region. We previously reported RNA-based single-nucleotide-specific programmable riboregulators (SNIPRs) that leverage small variations in hybridization energy stemming from the distinctions in the target RNA sequence^18^. However, the speed of SNIPRs is limited by the time needed to translated reporter proteins in the cell-free systems, causing reactions to take at least 30 minutes^18^. Strand-displacement probes have been used in MARVE to provide rapid colorimetric mutation detection based on enzymatic cleavage of urea, but these probes are limited to detecting a single target per reaction^19^. Therefore, there remains a need to develop rapid, low-cost, low-leakage, and multiplexed point-of-care diagnostic technologies for identification of single-nucleotide mutations.

Fluorescent RNA aptamers are a class of RNA molecules that bind to specific fluorogenic molecules to generate a fluorescent signal^20^. RNA aptamers are typically discovered through selection processes^20–22^. Many different types of fluorescent RNA aptamers have been selected with varying emission wavelengths and providing optimized quantum efficiencies^23–29^. Since the discovery of the fluorescent RNA aptamers^30^, they have been widely used in bioanalysis and bioimaging as fluorescent probes^31–34^. The most straightforward way to utilize a fluorescent RNA aptamer is by fusing its sequence to RNA targets as a reporter, enabling the detection of target RNA molecules either through fluorescent imaging or a fluorimeter^33–36^. Because the fluorescent RNA aptamer must fold into the correct structure to generate a fluorescent signal, an alternative approach involves designing a target-dependent molecular reconfiguration from a non-fluorescent state to a fluorescent state through splitting^31,32,37,38^ or refolding^39–41^ of the RNA aptamer. Recently, we reported aptamer-based switches that employ a toehold-mediated mechanism for rapid detection of different pathogens from clinical samples^41^. However, none of these methods can provide finer sequence information within a target RNA, such as identifying point mutations.

Herein, we introduce a class of fluorescent RNA switches based on fluorescent aptamers termed FARSIGHTs (for **<u>F</u>**ast **<u>A</u>**ptamer-based **<u>R</u>**eporter for **<u>SI</u>**ngle-nucleotide-specific **<u>G</u>**enotyping through **<u>H</u>**ybridiza**<u>T</u>**ion) that can identify single nucleotide mutations in an RNA target with high discrimination performance without the assistance of enzymes. The FARSIGHT design consists of two regions: the target recognition region and the fluorescent RNA aptamer configuration region.

In the absence of the mutation of interest in the RNA target, the RNA switch remains in its original state, and no fluorescent signal is generated. However, the target recognition region can identify single-nucleotide mutations in an enzyme-free manner and trigger reconfiguration of the RNA aptamer region through a stabilized basal stem, which refolds to bind to the small fluorogenic ligand molecule, releasing a fluorescent signal in as little as 5 min. This design is universal and can be applied to identify any mutation using different fluorescent RNA aptamers, such as Broccoli, Corn, and Red Broccoli, enabling multiplexed mutation identification. Moreover, the switch can be integrated with isothermal amplification reactions to identify mutations from clinical samples with sensitivity comparable to reverse transcription quantitative PCR (RT-qPCR). For whole RNA extracted from 23 clinical saliva samples, multiplexed assays using FARSIGHTs simultaneously distinguish SARS-CoV-2 Omicron variants from SARS-CoV-2 variants Alpha, Beta, and Gamma with an accuracy of 100% concordant with RT-qPCR and genomic sequencing. Overall, this study not only provides valuable tools for studying RNA structure and function but also opens up new avenues for designing programmable molecular devices with applications ranging from disease biomarker detection to advanced aptamer-based regulatory elements that operate in vivo.

## RESULTS

### FARSIGHT Design Principles

To implement aptamer-based systems with single-nucleotide specificity, we took advantage of two design principles exploited by previous highly specific detection systems. First, toehold exchange probes^42^ and related systems like SNIPR^18^ have achieved SNP detection by using strand-displacement reactions that operate near chemical equilibrium. These toehold exchange systems employ forward and reverse toehold regions of similar binding energies to enforce sequence-specific release of a single-stranded reporter when the correct sequence is detected (**Fig. 1a**). When the target sequence has a single-base difference, the resulting mismatch leads to a significant energy penalty that prevents release of the single-stranded domain. Second, studies with fluorescent RNA aptamers have demonstrated that the binding of fluorogenic molecules to the aptamer only occurs when it is properly folded with an intact basal stem^20,23,41^. Accordingly, formation of the aptamer basal stem can be used to turn on aptamer fluorescence when a target nucleic acid binds. To reconcile these two strategies, which require release of a single-stranded domain and formation of a stem structure, respectively, we adopted an alternative mechanism for FARSIGHTs that employs a domino-like intramolecular interaction to trigger aptamer folding.

**Fig. 1.**
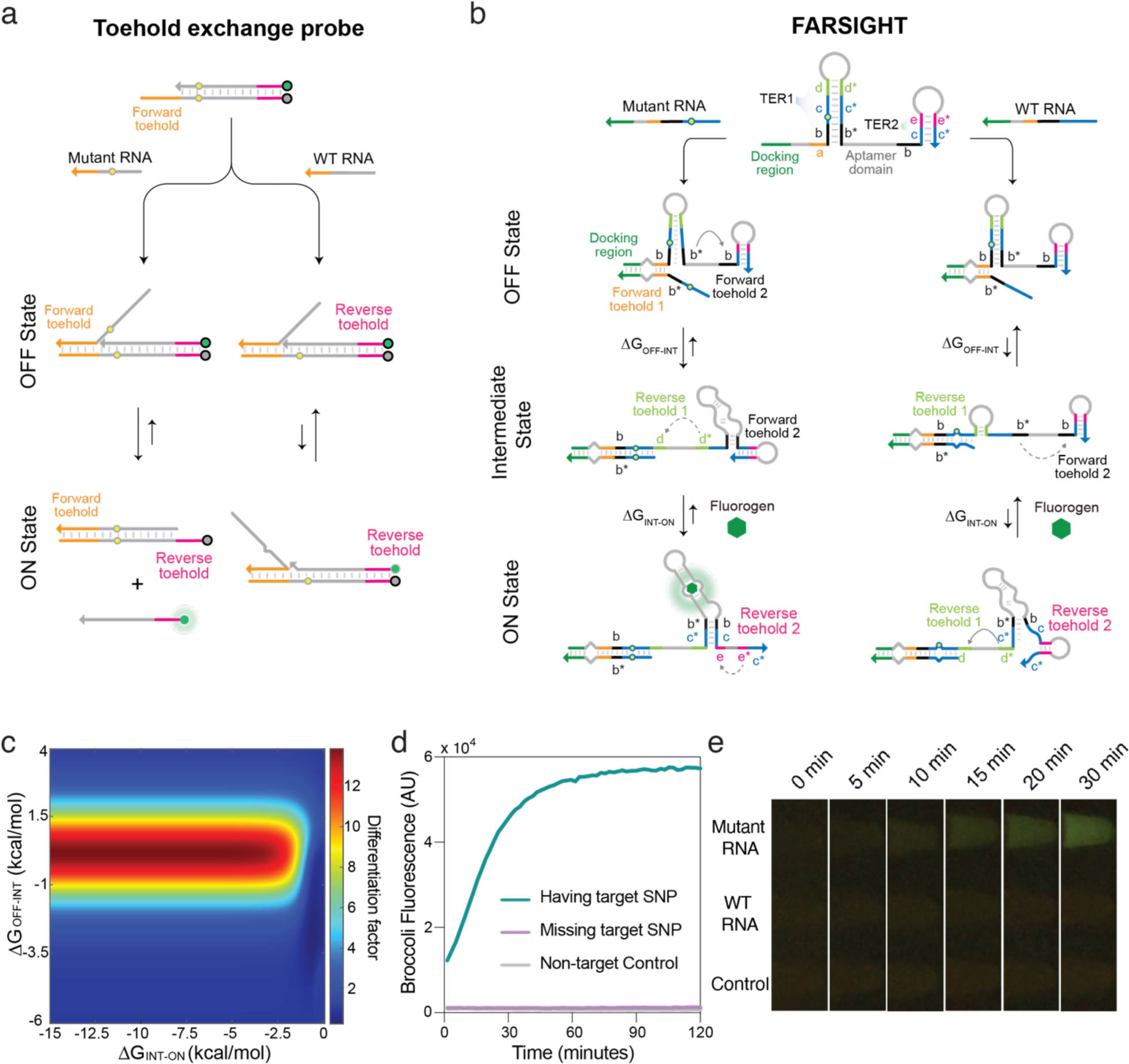
FARSIGHT operating mechanism and initial system characterization. **a**, Design schematic of a conventional toehold exchange probe that consists of two nucleic acid strands, the probe strand and the protector strand, that are hybridized to each other. The binding between the probe strand and the target strand results in the protector strand being released only if the target has the correct sequence. **b**, FARSIGHTs consist of (i) a target-recruiting docking site (shown in green) that is complementary to a target nucleic acid for promoting contact between RNA target and a hairpin structure sequestering the stabilizing stem (**c*-b\***) of the aptamer sequence; (ii) toehold exchange region 1 (TER1) having forward toehold 1 (**a**, shown in amber) and reverse toehold 1 (**d**, shown in lime); (iii) the core reporter aptamer sequence; and (iv) toehold exchange region 2 (TER2) having forward toehold 2 (**b**, shown in black) and reverse toehold 2 (**e**, shown in pink). The RNA target carrying the mutation of interest yields a favorable free energy in the reaction with the FARSIGHT, initiating a branch migration from forward toehold 1 that opens the hairpin stem. The newly released **c*-b\*** domain binds to the downstream **b-c** domain in TER2, enabling the fluorescent output from the aptamer/fluorogen complex (ON state). An incorrect target RNA without the expected mutation results in an energy penalty from the mismatched sequence and firmly shifts the equilibrium to OFF state. **c**, Biochemical modeling predicts the FARSIGHT design can detect point mutations in different ranges of ΔGOFF-INT and ΔGINT-ON. **d**, Time-course measurements of fluorescence from a FARSIGHT with a Broccoli reporter aptamer with correct or incorrect synthetic targets that have a single-nucleotide difference, or without a synthetic target. **e**, Photographs of fluorescence from FARSIGHT reactions for discriminating single-nucleotide mutations. Photographs were taken at 0, 5, 10, 15, 20, and 30 min of the reaction using a blue-light transilluminator.

As shown in **Fig. 1b**, the FARSIGHT design makes use of a pair of coupled toehold exchange reactions engineered that link RNA binding to aptamer stem formation. The two FARSIGHT toehold exchange reactions take place at two hairpin sites and make use of 3- to 6-nt forward and reverse toehold sites that can spontaneously hybridize or disassociate in typical reaction conditions. The two FARSIGHT hairpins also prevent folding of the reporter aptamer, resulting in minimal fluorescence in the absence of the target RNA. When the target first interacts with the FARSIGHT probe, it binds strongly to an extended ∼20-nt FARSIGHT docking region, previously used in SNIPRs^18^, providing it a stable site from which to initiate subsequent interactions as a quasi-unimolecular complex. In the first toehold exchange reaction, the target RNA binds to the forward toehold domain **a**, and the ensuing branch migration reaction disrupts the first FARSIGHT stem through to the reverse toehold domain **e**. This toehold domain opens spontaneously and thus completes the exchange reaction when a target with the correct sequence binds. Completion of this first strand-displacement rection establishes the intermediate state of the FARSIGHT-target complex, and the transition energy (ΔG_OFF-INT_) of the reaction is controlled by the forward toehold domain **a** and reverse toehold domain **d**. The newly released single-stranded domains **c\*** and **b\*** from this reaction are then free to initiate the second toehold exchange reaction with the forward toehold domain **b** in the second FARSIGHT stem region. The ensuing branch migration reaction proceeds until it reaches the second reverse toehold domain **e**, which unwinds spontaneously to complete the reaction. The second strand-displacement rection yields the ON state of the FARSIGHT-target complex with the transition energy (ΔG_INT-ON_) of the reaction controlled by the forward toehold domain **b** and reverse toehold domain **e**. Importantly, the second toehold exchange process leads to formation of the aptamer basal stem, which promotes proper aptamer folding. Once the reporter aptamer folds, it binds to the cognate fluorogen to generate a fluorescent readout signal.

From this base design, FARSIGHTs must satisfy three key rules to reliably identify point mutations: (1) the toehold and branch migration domains of the first toehold exchange reaction must perfectly match with the cognate region of the mutant target, (2) the equilibrium energy between the OFF state and intermediate state ΔG_OFF-INT_ should be designed to be around −1 kcal/mol, and (3) the equilibrium energy between the intermediate state and the ON state ΔG_INT-ON_ should also be designed to be −1 kcal/mol or lower. Therefore, the first strand displacement reaction is a slightly thermodynamically favorable step, and the intermediate state will be the dominant state upon the binding of mutant target with the correct sequence. However, in the presence of the WT target, which does not have the correct sequence, the resulting mismatch will leave a single-nucleotide bulge in the intermediate state (**Fig. 1b**, **Supplementary Table 1**). The bulge will cause a ∼4 kcal/mol energy penalty and shift the equilibrium energy of the transition between the OFF state to intermediate state to ∼3 kcal/mol. Consequently, the first toehold exchange reaction becomes unfavorable upon the binding of WT targets. In the event that the first reaction proceeds, the second toehold exchange reaction provides a final check on sensor activation through two mechanisms. First, it impedes fluorogen-driven aptamer folding, which would otherwise contribute to signal leakage. Second, it delays aptamer folding and thus provides additional time to reverse the first strand displacement for incorrect targets. Overall, the use of the coupled toehold exchange reactions enables us to tune the favorability of the sensor activation steps depending on the point mutation of interest in the RNA target and the aptamer/fluorogen pair.

We developed a biochemical model to capture the key steps of the FARSIGHT performance, including the first and the second step strand displacement with the binding of fluorogenic molecules (see **Supplementary Note 1** and **Supplementary Fig. 1-2** for details). A combination of different chemical equilibrium energies for the two-step strand-displacement reaction has been systematically explored. As shown in **Fig. 1c**, the optimal FARSIGHT design will have an equilibrium energy ranging from −2 to 2 kcal/mol for the transition from OFF state to Intermediate state and an equilibrium energy ranging from −1.5 to −15 kcal/mol for the second transition from Intermediate state to ON state. The energy of this second transition does not include the binding energy of the fluorogen to the RNA aptamer.

### Detection of mutations using FARSIGHTs

We designed an initial FARSIGHT with the Broccoli aptamer and targeted it to the P681R mutation in the SARS-CoV-2 S gene based on the principles described above. The reaction energies for the two toehold exchange reactions are −1.9 kcal/mol and −1.8 kcal/mol, respectively. Well-folded Broccoli aptamer binds to its cognate fluorogen DFHBI-1T with an equilibrium energy of −8.87 kcal/mol (see **Supplementary Note 2** for binding analysis), which provides a strong green emission signal under blue-light illumination^23^. We then experimentally validated the mutation identification capability of the FARSIGHT design. We in vitro transcribed the FARSIGHT RNAs and incubated them with DFHBI-1T. We found that no fluorescent signal was generated. The addition of mutant RNA target into the incubated mixture gave very strong fluorescent signal within 30 minutes, while the addition of WT target yielded minimal fluorescence compared to the control. A typical kinetic curve and fluorescence images upon the blue light illumination are shown in **Fig. 1d-e** (**Supplementary Table 1**, **Supplementary Video**).

To systematically study how the equilibrium energy influences FARSIGHT discrimination capability, we designed a set of 99 (see **Supplementary Table 1**) different FARSIGHT RNAs by varying the binding strength of the four different toehold domains. The results are shown in the heatmap in **Fig. 2a** and demonstrate good agreement with the biochemical modeling results in **Fig. 1c**. The small discrepancy between the simulation results and experimental results (∼1 kcal/mol between the ΔG_OFF-INT_) may be attributed to two major factors: (1) inaccuracies in predictions of RNA folding by current RNA free energy prediction models^43^ and (2) difficulties in accurately representing the intermediate state, which is highly dynamic and has multiple possible structures depending on the FARSIGHT sequence. For example, we observed two designs that yielded identical equilibrium energies for both toehold exchange reactions but provided dramatically different differentiation factors in experiments (**Supplementary Fig. 3** and **Supplementary Table 1**). Despite these limitations, the model can assist in the design of functional FARESIGHTs to target different mutations in an RNA transcript.

**Fig. 2.**
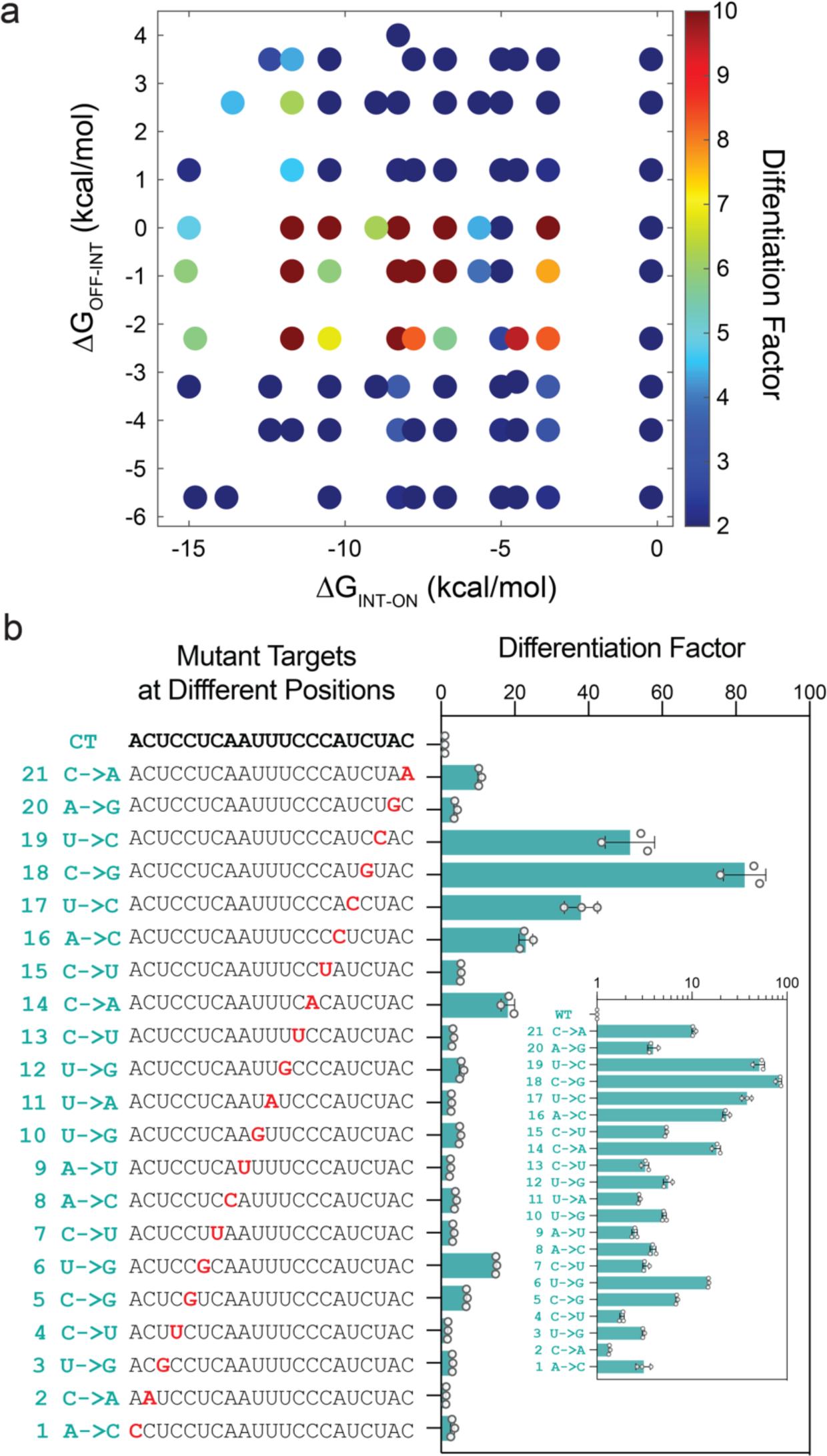
FARSIGHT discrimination capability of mutations by equilibrium energy. **a**, The differentiation factor of FARSIGHTs with varying binding strength of the four different toehold domains upon binding to the mutant targets. Reactions were measured in triplicate. **b**, Differentiation factor obtained after a 2 h reaction for each mutated target RNA with single-nucleotide substitutions including G-U wobble base pairs enables all of them to be distinguished from the correct target sequence. Inset: Differentiation factor measured for Broccoli FARSIGHTs on a logarithmic scale. Relative errors for the differentiation factor were obtained by adding the relative errors of the FARSIGHT fluorescence measurements with the correct target and the SNP-containing target in quadrature. Relative errors for the correct and SNP-containing target sequence are from the s.d. of *n=*3 technical replicates.

We further evaluated the capacity of FARSIGHTs to discriminate mutations by examining mutations located at positions 1 to 21 in the target RNA (**Fig. 2b**, **Supplementary Table 2**), which hybridize from the beginning of the forward toehold to the end of the first branch migration domain. The RNA binding of this region affects the equilibrium between different states. To assess sensor performance, we employed the discrimination factor (Df), which is defined as the ratio of FARSIGHT fluorescence with the correct target divided by FARSIGHT fluorescence for the incorrect target. This set of targets revealed that the FARSIGHT probe could discriminate mutations at all 21 target positions. We also found that mutations that convert canonical Watson-Crick base pairing to G-U wobble base paring can also be distinguished, such as the A->G mutation at position 20 and C->U at position 15. However, mutations at positions 16 to 19 yielded the best discrimination results, providing discrimination factors all above 50. Although mutations at either the toehold or branch migration domain will have the same influence on the chemical equilibrium energy, the improved specificity at positions 16 to 19 suggests additional contributions from kinetic effects. In particular, mutations at positions 16 to 19, which displace the FARSIGHT b* domain, likely provide a stronger kinetic trap to prevent FARSIGHT activation for mutations in this region by inhibiting release of the toehold-binding domain for the second strand-displacement step.

### Automated in silico Design of FARSIGHTs and Rapid Validation in vitro

To test the universality of FARSIGHTs for clinically relevant mutations, we generated a set of FARSIGHTs to target critical mutations in the SARS-CoV-2 RNA genome (**Fig. 3a**, **Supplementary Table 3**) using computational design (see **Supplementary Note 2**). We selected mutations that defined different SARS-CoV-2 variant strains that evolved during the COVID-19 pandemic, including Alpha (B.1.1.7), Beta (B.1.351), Gamma (P.1), Delta (B.1.617.2), and Omicron (B.1.1.529) as well as its sublineages (see **Supplementary Fig. 4a**). The resulting FARSIGHT transcripts were then systematically evaluated for specificity by challenging them with synthetic versions of the SARS-CoV-2 wild-type or mutant sequences after in vitro transcription. The discrimination factor was determined for each of these FARSIGHTs to quantitate their discriminatory capacity.

**Fig. 3.**
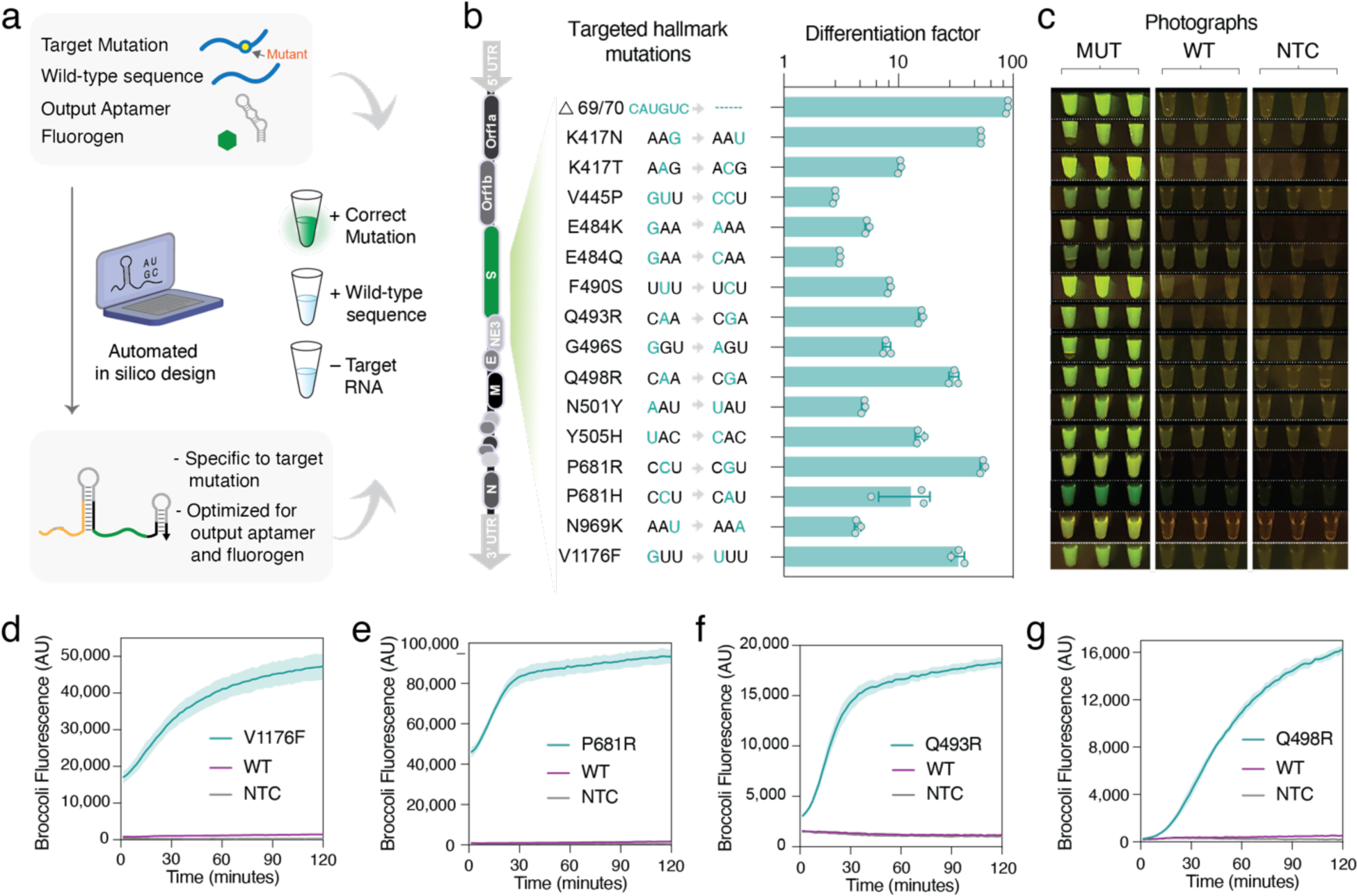
*In silico* design and discrimination of SARS-CoV-2 mutations by FARSIGHTs with Broccoli output. **a**, The in silico FARSIGHT design process takes the mutant target sequence to be detected, the wild-type sequence to be avoided, and the desired aptamer/fluorogen output to generate candidate FARSIGHTs that are then tested in vitro. **b**, Differentiation factors obtained after 2 h reactions for optimal Broccoli FARSIGHTs determined in the presence of the correct variant sequence or the wild-type sequence. Individual points show the fold fluorescence increase from *n=*3 technical replicates. Relative errors for the differentiation factors were obtained by adding the relative errors of the ON and OFF state fluorescence measurements in quadrature. **c**, Photographs of FARSIGHTs in **b** after 2 h reactions using a blue-light transilluminator. **d**-**g**, Time-course measurements of the fluorescence signal from top-performing Broccoli FARSIGHTs in the presence of the correct mutant variant or wild-type targets. Shaded regions denote mean ± s.d. with *n*=3 technical replicates.

**Figure 3b** shows the discrimination performance of the systems using Broccoli as the output aptamer. From these rapid screening experiments (**Supplementary Fig. 4b** and **Supplementary Table 4**), we find that all target mutants can be distinguished from the wild-type sequences to facilitate the recognition of viral variants (**Supplementary Fig. 5a** and **Supplementary Table 4**). Out of this set, the top-performing FARSIGHT provided a large ≥90-fold increase in fluorescence upon discrimination of the cognate mutant compared to reactions with wild-type sequence, and half of the FARSIGHTs yielded discrimination factors of at least 10.

Clearly visible is the strong green fluorescence produced by the Broccoli FARSIGHTs containing the SARS-CoV-2 target RNA carrying the targeted mutations (**Fig. 3c**). **Figure 3d-g** and **Supplementary Fig. 5b-I** show time-course measurements of the sensors profiling activation speed and leakage of the sensors in 37°C reactions. Statistically significant fluorescence signals can be detected after 5 minutes of reaction in a plate reader for the fastest sensors and strong activation is observed within 30 minutes for the optimal Broccoli FARSIGHT designs. Impressively, for the reactions with wild-type RNA without the targeted mutations, only near-background fluorescence could be observed in many of the sensors.

### Multiplexed detection of point mutations using FARSIGHTs with different aptamer outputs

The rapid and accurate identification of multiple mutations in a single test can be especially useful in monitoring the evolution and dynamics of viral populations and help healthcare professionals make appropriate treatment decisions. The design of FARSIGHTs, as described here, can be adapted for use with a wide range of fluorogenic aptamers, including but not limited to Broccoli, Corn, and Red Broccoli, which could be used to increase assay multiplexing capacity or allow it to be interpreted by other fluorescence detection systems (**Fig. 4a**). Red Broccoli and Corn FARSIGHTs were designed for targeting a series of mutations characteristic of SARS-CoV-2 variants (**Supplementary Fig. 6-7**, **Supplementary Table 5**-**6**). **Figure 4b-c** shows the performance evaluation for a library of 7 Red Broccoli FARSIGHTs targeting the sense orientation or antisense orientation of the mutant target sequence. Red Broccoli FARSIGHTs exhibited differentiation factors of over 10 for most of the mutant targets with a very low level of leakage and fast activation time with strong fluorescence occurring within 15 minutes (**Fig. 4c**, **Supplementary Fig. 6b-g**). The FARSIGHT designs with Corn were also successful in detecting different target mutations as shown in **Fig. 4d-e** and **Supplementary Fig. 7**. These Corn FARSIGHTs display lower ON/OFF ratios compared to the Broccoli-based systems as they emit relatively weaker fluorescence output with a weaker reliance on a strong basal stem^41^.

**Fig. 4.**
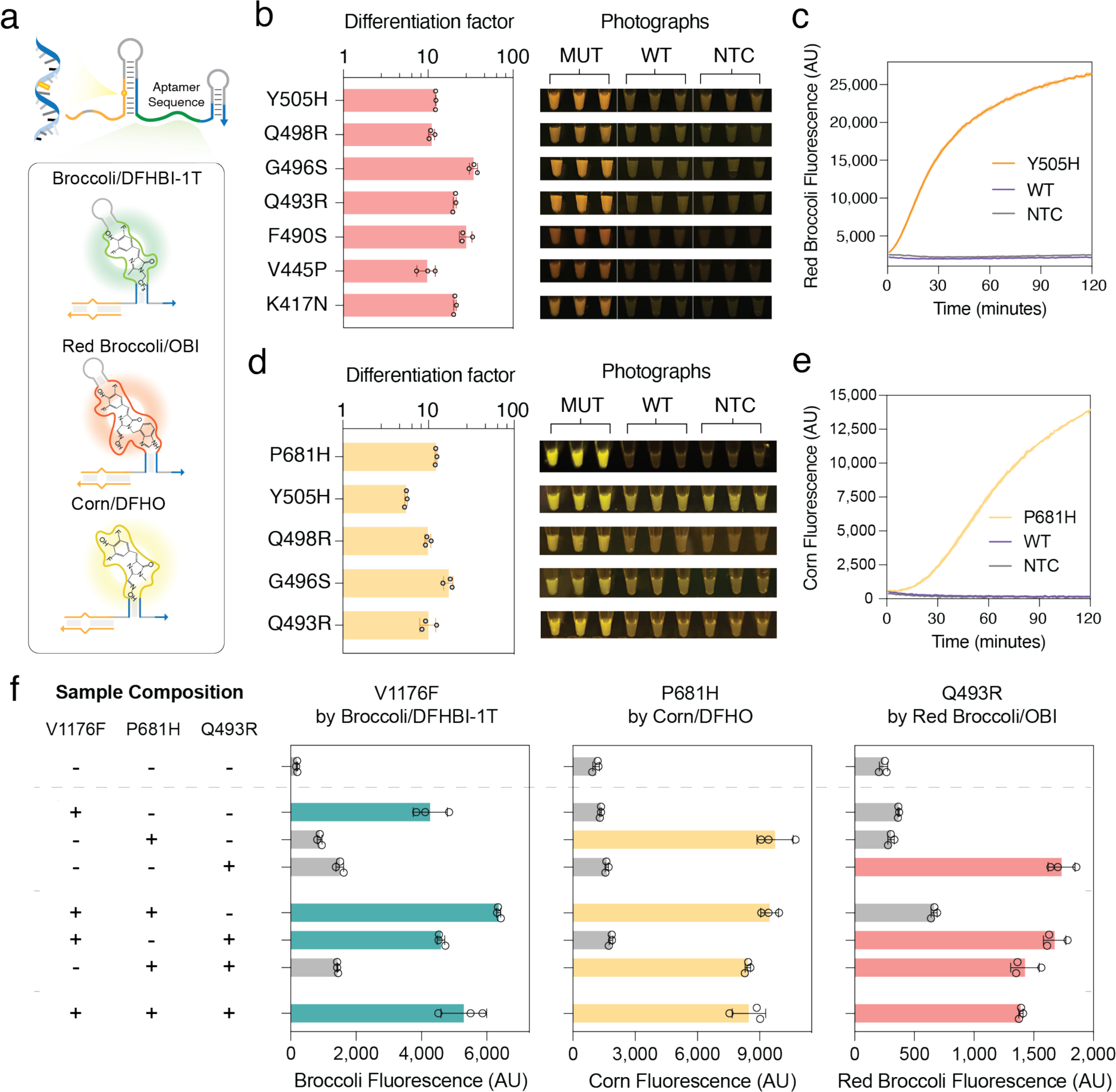
Multiplexed detection of single point mutations by orthogonal FARSIGHTs. **a**, Schematic of the FARSIGHT design with different output aptamer/fluorogen combinations for multiplexed mutation detection. Each aptamer has its own sensor RNA that can be independently activated by its correct mutation-carrying RNA target and induce folding of a distinct aptamer reporter. **b**, Representative differentiation factor and visual detection of fluorescence from FARSIGHTs with Red Broccoli/OBI output. Differentiation factors represent mean fluorescence fold increase of the correct mutant target over the wild-type sequence without the target SNP after 2 h reactions. Relative errors for fold increases were obtained by adding the relative fluorescence errors in quadrature for *n=*3 technical replicates. Photographs were taken using a blue-light transilluminator. **c**, Time-course measurements of fluorescence from the Y505H-targeted Red Broccoli FARSIGHT with correct mutant target or wild-type sequence and without target RNA. Shaded regions denote mean ± s.d. of *n*=3 technical replicates. **d**, Representative differentiation factor and visual detection of fluorescence from FARSIGHTs with Corn/DFHO output. Differentiation factors represent logarithmic mean fluorescence fold increase of the correct mutant target over the wild-type sequence without the target SNP after 2 hr reactions. Relative errors for fold increases were obtained by adding the relative fluorescence errors in quadrature for *n=*3 technical replicates. Photographs were taken using a blue-light transilluminator. **e**, Time-course measurements of fluorescence from the P681H-targeted Corn FARSIGHT with the correct mutant target or wild-type sequence and without target RNA. Shaded regions denote mean ± s.d. with *n*=3 technical replicates. **f**, Simultaneous multiplexed detection of V1176F, P681H, and Q493R using FARSIGHTs with Broccoli/DFHBI-1T, Corn/DFHO, and Red Broccoli/OBI reporters, respectively. Samples with or without those mutations are applied for a 2 hr FARSIGHT reaction to test the multiplexed mutation analysis performance. Bars denote the mean ± s.d. of *n=*3 technical replicates.

We then moved on to differentiate multiple mutations with three aptamers having spectrally distinct fluorescence. The V1176F (for characterizing Gamma) and P681H (for characterizing Alpha) S gene mutations were detected using orthogonal FARSIGHTs with Broccoli/DFHBI-1T and Corn/DFHO outputs, respectively, in the same reaction (**Supplementary Fig. 8a**, **Supplementary Table 7**). Similarly, as shown in **Supplementary Fig. 8b**, the orthogonal FARSIGHTs with Broccoli/DFHBI-1T and Red Broccoli/OBI outputs also recognized all combinations of N969K (for all Omicron variants) and G496S (for Omicron BA.1 sub-variant) S gene mutations. For triplex detection, all combinations of V1176F, P681H, and Q493R S gene mutations were successfully detected by FARSIGHTs programmed with Broccoli/DFHBI-1T, Corn/DFHO, and Red Broccoli/OBI, respectively (**Fig. 4f**, **Supplementary Table 7**).

### Integration with isothermal amplification for low concentration RNA targets

Accurate identification of viral variants of interest from other variants is crucial for the early detection and management of emerging outbreaks. We coupled FARSIGHTs with nucleic acid sequence-based amplification (NASBA) reactions^44^, which enable nucleic acid amplification at a constant 37°C to 42°C temperature for detecting clinically relevant RNA concentrations^45^ (**Fig. 5a**). NASBA begins with a reverse primer that facilitates reverse transcription of the target RNA, forming an RNA/DNA duplex. RNase H degrades the RNA template, allowing a forward primer with a T7 promoter to bind and initiate elongation of the complementary strand, creating a double-stranded DNA product. T7-mediated transcription of the DNA template generates multiple copies of the target RNA sequence^44,46^.

**Fig. 5.**
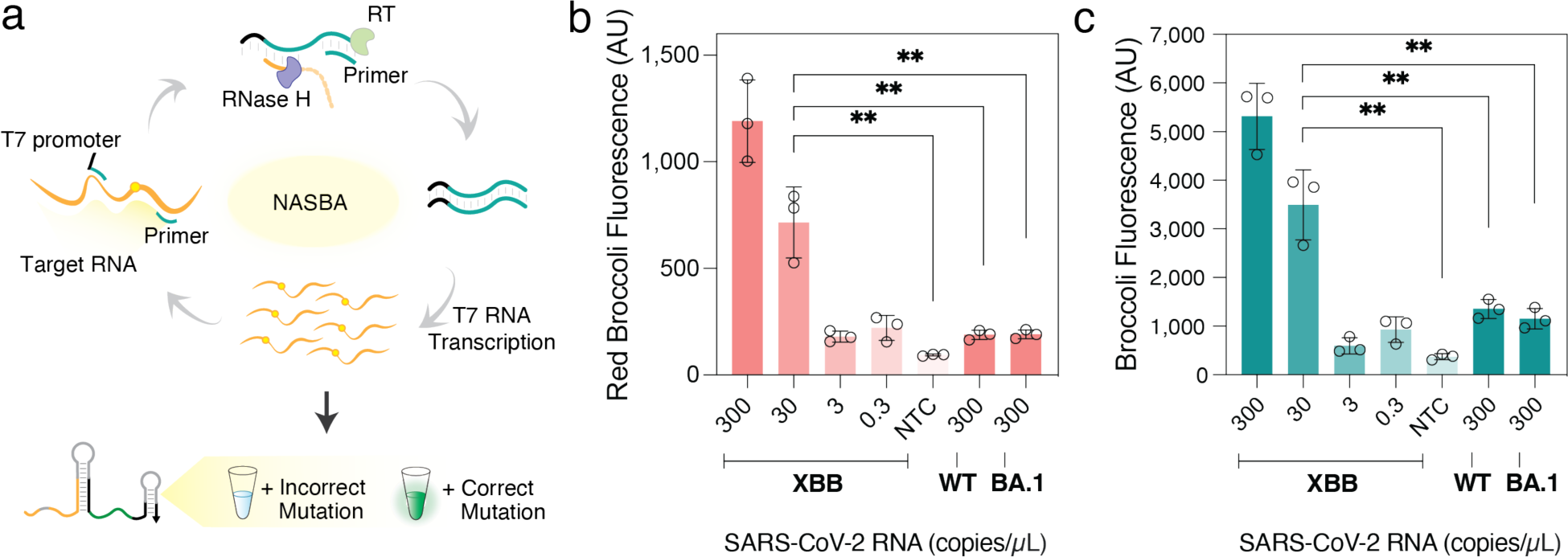
Coupling Isothermal amplification with FARSIGHTs to identify SARS-CoV-2 variants. **a**, Schematic of assay workflow for the identification of SARS-CoV-2 variants. An RNA sample is first amplified using NASBA (nucleic acid sequence-based amplification). The resulting amplified RNA with the correct target mutant activates the FARSIGHT to produce an output fluorescence signal to uncover the viral variant. **b**-**c**, Determination of the detection limit of FARSIGHTs for the F490S S gene mutation of the XBB Omicron subvariant in SARS-CoV-2 RNA through Red Broccoli/OBI (**b**) and Broccoli/DFHBI-1T reporters (**c**), respectively. The signal of samples at different concentrations was obtained as the mean fluorescence intensity ± s.d. of FARSIGHTs at 2 h, which was preceded by an 80-minute NASBA reaction. (n = 3 technical replicates; bars represent arithmetic mean ± s.d; two-tailed Student’s t test; ns, p > 0.05; **, p < 0.01).

NASBA primer sets for the SARS-CoV-2 RNA genome were designed and screened experimentally (**Supplementary Fig. 9**, **Supplementary Table 8**). The optimal primer pairs were then combined with the top-performing FARSIGHTs from screening tests for a series of experiments supplying 80-minute amplification reactions with different concentrations of cultured variant SARS-CoV-2 RNA containing either the target mutation or WT SARS-CoV-2 RNA (**Fig. 5**, **Supplementary Fig. 10**, **Supplementary Table 9**). We designed the FARSIGHT to target the F490S S gene mutation^47,48^ to identify the XBB Omicron subvariant of SARS-COV-2. As shown in **Fig. 5b-c**, upon coupling with NASBA, the Red Broccoli and Broccoli FARSIGHTs targeting F490S displayed a substantial fluorescence increase for XBB RNA template concentrations down to 30 RNA copies/µL, yet the reactions utilizing cultured wild-type or Omicron BA.1 SARS-CoV-2 variants only provided low fluorescence. The detection limits achieved using our approach are sufficient for detection of the virus from clinical samples^49,50^.

### SNP-identification of SARS-CoV-2 variants in clinical saliva samples

We then applied the NASBA-integrated FARSIGHT assay to detect the SARS-CoV-2 virus and further identify the specific variant in clinical saliva samples. We used two designed FARSIGHTs to detect the N501Y and Y505H mutations with Broccoli and Red Broccoli (**Fig. 6a**, **Supplementary Table 9**), respectively. The mutation N501Y exists in all the major SARS-CoV-2 variaints^51^, and Y505H is a hallmark mutation for the Omicron variant^52,53^. Synthetic virus RNA fragments from Omicron and Alpha variants were first used to validate the two FARSIGHTs. We used the 95^th^ percentile values of 33 Alpha RNA samples and 35 SARS-CoV-2 negative samples to determine the threshold value (arbitrary units (AU)) for identifying Omicron variants (882.5 AU from Red Broccoli FARSIGHTs) and general SARS-CoV-2 variants (2329.7 AU from Broccoli FARSIGHTs), respectively (**Supplementary Fig. 12a**). As shown in **Fig. 6b**, the Omicron RNA genome fragments can turn on the fluorescence signal in both green and red channels for the two FARSIGHTs, resulting in orange output fluorescence. The Alpha variant can only activate the fluorescent signal in the green channel from the FARSIGHT targeting N501Y. Similarly, this strategy can also be adopted to identify XBB subvariant (F490S mutation) from general Omicron variants (Y505H mutation) or identify the Omicron BA.1 variant (G496S) from major SARS-CoV-2 (N501Y mutation) variants (**Supplementary Fig. 11**, **Supplementary Table 10**).

**Fig. 6.**
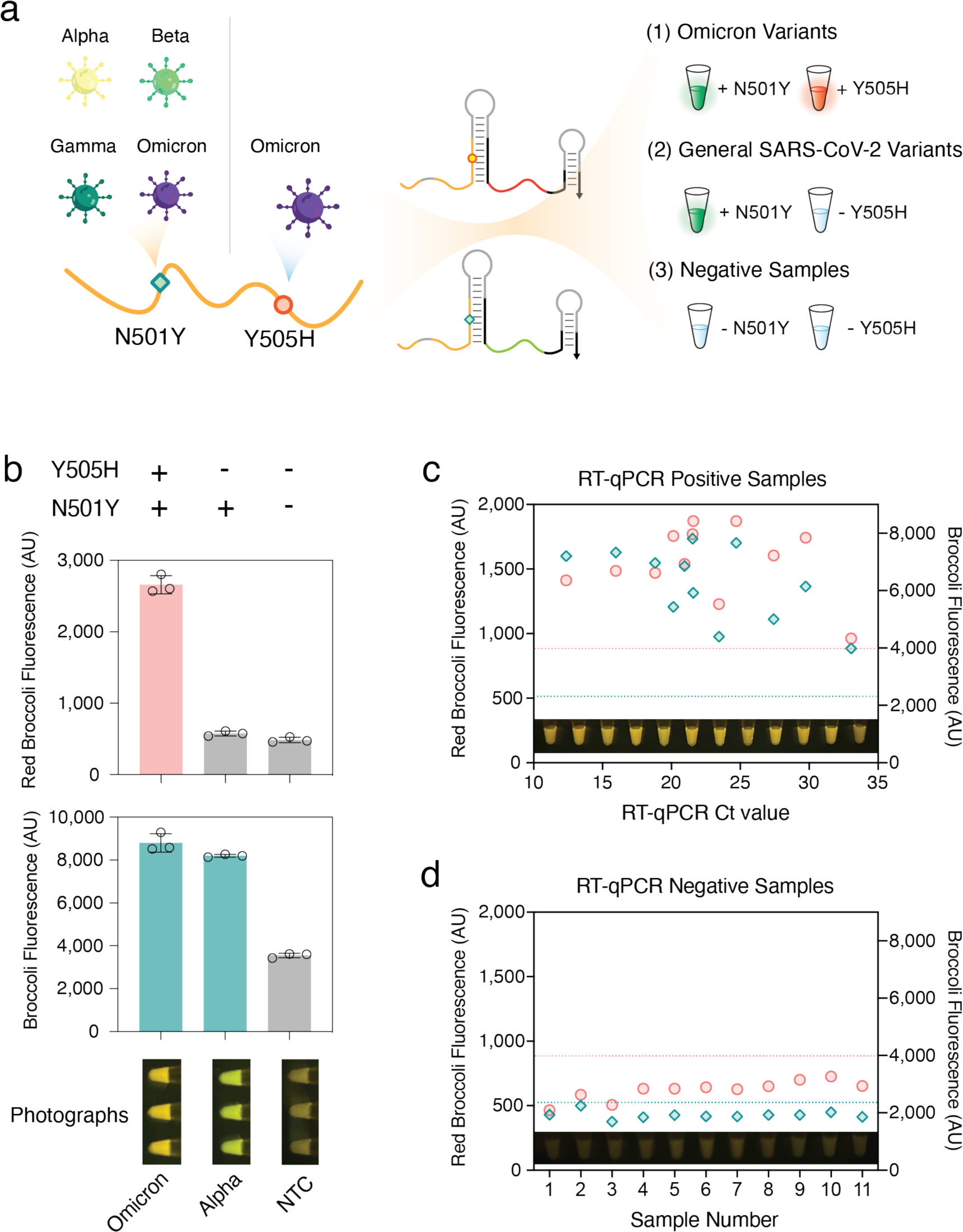
Identification of SARS-CoV-2 variants from clinical saliva samples using FARSIGHTs. **a**, Schematic for simultaneous detection of two mutation hotspots in the SARS-CoV-2 RNA genome to identify virus strains from patient saliva samples. A rapid increase in the fluorescence of Red Broccoli and/or Broccoli FARSIGHTs signifies the presence of the Omicron variant and general SARS-CoV-2 variants (Alpha, Beta, Gamma, and Omicron), respectively. **b**, Red Broccoli FARSIGHTs distinguish Omicron variants from others by detecting the Y505H S gene mutation. General SARS-CoV-2 variants are identified by N501Y-targeted Broccoli FARSIGHTs. (*n=*3 technical replicates; bars represent the arithmetic mean ± s.d.) **c**-**d**, Fluorescence signal obtained from 12 clinical positive saliva samples with Omicron BA.5 and 11 negative clinical saliva samples after 2 h FARSIGHT reactions (n=3 technical replicates, bars represent the arithmetic mean ± s.d.). Coral circles correspond to Red Broccoli FARSIGHT fluorescence for the Y505H S gene mutation and teal diamonds correspond to Broccoli FARSIGHT fluorescence for the N501Y S gene mutation. The dashed lines represent theoretical diagnostic thresholds that were determined using 95^th^ percentile fluorescence values for non-template controls of Omicron and general SARS-CoV-2 strains (coral dash line: Red Broccoli FARSIGHT threshold value for determining Omicron strains by identifying the Y505H S gene mutation; teal dashed line: Broccoli FARSIGHT threshold value for determining general SARS-CoV-2 variants by identifying the N501Y S gene mutation).

We next validated the FARSIGHT dual-site discrimination assay using clinical saliva samples from 12 positive and 11 negative patients and compared with RT-qPCR test. The viral RNA sample was extracted from the saliva and applied to the NASBA-coupled FARSIGHT assay. All the positive patient samples gave a strong fluorescent signal increase in both green and red channels (**Fig. 6c**, **Supplementary Fig. 13**), and negative samples did not show significant signal change in either channel (**Fig. 6d**, **Supplementary Fig. 14**). These results indicate that all the positive patients are infected by the Omicron variants, aligning well with the dominance of the Omicron variant in the US when the samples were collected. The RT-qPCR and FARSIGHT results for the samples matched very well displaying complete concordance (**Supplementary Fig. 12b**).

## DISCUSSION

We have developed FARSIGHTs, RNA switches that couple fluorescent aptamer-based readout with robust single-nucleotide mutation discrimination capabilities. FARSIGHTs exploit a coupled pair of toehold exchange reactions that operate at exquisitely designed chemical equilibrium and achieve high sequence specificity. We have demonstrated that FARSIGHTs are compatible with multiple fluorescently distinct aptamers, enabling one-pot multiplexed mutation detection, and can successfully identify arbitrary mutations, including those generating G-U wobbles. Through integration with isothermal amplification reactions, FARSIGHTs can detect mutations in low-abundance RNA targets extracted from clinical saliva samples. When paired with a portable fluorescent reader and lyophilization of the necessary biological reagents, FARSIGHTs hold significant promise as a convenient diagnostic system for detecting mutations in diverse settings, including homes, hospitals, and field environments.

Compared with RNA sequencing^54,55^ and PCR-based^56^ methods commonly employed in central laboratories, FARSIGHTs require a significantly lower cost of ∼$1.76 per reaction (see **Supplementary Note 3**), a simpler equipment setup, and provide a faster turnaround time without requiring companion enzymes for readout. Moreover, they have several advantages over recently developed mutation detection methods. For instance, use of CRISPR-Cas enzymes for mutation detection necessitates a protospacer adjacent motif (PAM) sequence in the double-stranded DNA target or provides point mutation detection in only a small sequence region in single-stranded targets based on the seed region of the spacer^57,58^, while FARSIGHTs can resolve point mutations that occur over a 21-nt window. In comparison to FARSIGHTs, SNIPRs can generate colorimetric outputs, but they have slower reaction times and require more sophisticated and costly cell-free expression systems to generate signal because of their translation-based output^18,59^. Similarly, the utilization of conventional toehold exchange probes demands costly modifications involving dyes and quenchers for each target, thus increasing development and deployment costs. Although the use of unmodified toehold exchange DNA probes in MARVE decreases its cost^19^, the colorimetric readout of MARVE relies on urease activity and is incompatible with one-pot multiplexing.

The design of FARSIGHTs provides new insight into the integration of RNA folding control with its biological roles. RNA’s function is related to its diverse structures, and the design of functional RNA molecular devices thus demands control of RNA folding for a variety of different structure types. Conventional single-step strand-displacement reactions are aptly suited for release of single-stranded RNA functional motifs. Indeed, this approach has been used to control gene translation^18,60–62^, transcription^63^, and genome editing^64,65^. However, there are many cases where a functional RNA must fold into fully or partially double-stranded states to operate, such as aptamers^23–25,27,41,66^, ribozymes^67–69^, tRNAs^70,71^, and microRNA precursors^72,73^. Using the domino-like toehold exchange reactions of FARSIGHTs, we expect that it will be possible to not only regulate the function of stem-containing functional RNAs but to make them responsive to minute sequence differences in target transcripts.

Since the FARSIGHT mechanism relies on small differences in folding free energy, it is possible that a variety of other target molecules could be detected using similar strategies based on intramolecular refolding. For instance, RNA modifications are known to affect the binding affinity of RNAs^18,74^ and could thus be differentiated by FARSIGHTs. Furthermore, FARSIGHT-based detection could be extended to other types of molecules, such as small molecules^75,76^ or proteins^77–79^, when their RNA binding motif is embedded into the first step of strand displacement to shift chemical equilibrium to favor aptamer folding. FARSIGHTs also have great potential to visualize mutations in live cells. Fluorescent RNA aptamers have been widely used for live-cell RNA imaging^32,80,81^, but these aptamer-based probes have not demonstrated single-nucleotide specificity for spatial profiling at the single-cell level. FARSIGHTs that operate in live cells could allow us to study the occurrence of genome mutation in real time, providing deeper spatial and temporal information into biological processes such as DNA damage and repair^82,83^. We thus envision that FARSIGHTs and systems with similar mechanisms will provide valuable new insights into cell biology and wide biomedical applications in the future.

## MATERIALS AND METHODS

### FARSIGHT preparation

All DNA oligonucleotides were designed using the NUPACK software package as briefly described in the Results section and purchased from IDT (Integrated DNA technologies, Inc.). DNA fragments were assembled from 1 or 2 T7 promotor-having ssDNA templates with a T_m_ = 57 °C overlap region via PCR using Phusion High-Fidelity PCR Master Mix with HF Buffer (NEB, M0531L). FARSIGHT RNAs were transcribed from assembled PCR products (200 nM) in vitro using AmpliScribe™ T7-Flash™ Transcription Kits (Lucigen, ASF3507) from the DNA template at 37 °C for 4 hours. All transcription products were used without further purification unless otherwise noted.

### RNA target preparation

Synthetic target RNA for FARSIGHT *in vitro* validation experiments was directly transcribed from PCR-amplified product and was purified using Monarch® RNA Cleanup Kit (NEB, T2040L). RNA concentration was then determined by measuring the absorbance at a wavelength of 260 nm. The concentration for the synthetic target applied to the reactions was about 2 µM.

Full-genome synthetic SARS-CoV-2 variant RNA (controls 15, 16, 17, 23, 48 and 50) for other experiments (e.g. LoD tests, two-site simultaneous detection, and clinical samples validations) were purchased from Twist Bioscience. Full-genome reference SARS-CoV-2 RNA from heat-inactivated culture virions (VR-1986HK^TM^, Wild-type sequences) were purchased from ATCC.

### Readout of FARSIGHT reactions

A BioTek Synergy Neo2 multimode microplate reader (Agilent, NEO2MALPHAT) was used for all plate reader measurements. 1 μL direct FARSIGHT transcription product and 2.5 μM of purified target RNA was added to a 384-well plate along with 4 μM of DFHBI-1T for Broccoli (Lucerna, 410), 2 μM of DFHO for Corn (Lucerna, 500), or 4 μM of OBI for Red Broccoli (Lucerna, 610). DFHBI-1T, DFHO, and OBI buffers consist of 40 mM HEPES (Gibco™, 15630080), pH 7.4, 100 mM KCl (Invitrogen™, AM9640G), and 5 mM MgCl_2_ (Invitrogen™, AM9530G). Before each measurement, samples were shaken linearly for 30 seconds to ensure proper mixing. The plate reader was pre-heated, and the measurements were taken at 37°C unless otherwise noted. Fluorescence intensity values were corrected by subtracting the background fluorescence obtained from a blank control sample containing the FARSIGHT reaction without any RNA or DNA.

### Imaging of FARSIGHT reactions

To observe the fluorescence of the sensors, we illuminated the reactions in a microplate using a Safe Imager 2.0 Blue Light Transilluminator (Thermo Fisher, G6600). Videos and images of fluorescent reaction were taken by a standard cellphone camera (iPhone 14 Pro or iPhone 12, Apple Corp.).

### In-sample multiplexed point mutation tests

The two-input reactions with Corn and Broccoli outputs were prepared with 2 μ L Corn FARSIGHT/1.7 μM DFHO fluorogen and 1 μL Broccoli FARSIGHT/4 μM DFHBI-1T fluorogen. The 10X fluorogenic dye mix consisted of 17 μ M DFHO (Lucerna, 500), 40 μ M DFHBI-1T (Lucerna, 410), 40 mM HEPES (Gibco™, 15630080), pH 7.4, 100 mM KCl (Invitrogen™, AM9640G), and 5 mM MgCl_2_ (Invitrogen™, AM9530G).

The two-input reactions with Red Broccoli and Broccoli outputs were prepared with 1 μL Red Broccoli FARSIGHT/4 μM OBI fluorogen and 1 μL Broccoli FARSIGHT/4 μM DFHBI-1T fluorogen. The 10X fluorogenic dye mix consisted of 40 μM OBI (Lucerna, 610), 40 μM DFHBI-1T (Lucerna, 410), 40 mM HEPES (Gibco™, 15630080), pH 7.4, 100 mM KCl (Invitrogen™, AM9640G), and 5 mM MgCl_2_ (Invitrogen™, AM9530G).

The three-channel reactions were prepared with 2 μL Corn FARSIGHT/2 μM DFHO fluorogen, 1 μL Red Broccoli FARSIGHT/4 μM OBI fluorogen and 1 μL Broccoli FARSIGHT/4 μM DFHBI-1T fluorogen. The 10X fluorogenic dye mix consisted of 20 μM DFHO (Lucerna, 500), 40 μM DFHBI-1T (Lucerna, 410), 20 μM OBI (Lucerna, 610), 40 mM HEPES (Gibco™, 15630080), pH 7.4, 100 mM KCl (Invitrogen™, AM9640G), and 5 mM MgCl_2_ (Invitrogen™, AM9530G).

### NASBA/FARSIGHT reactions

10 μL NASBA reactions^44^ were carried out using the following protocol: 3.35 μL reaction buffer (Life Sciences, NECB-24; 3X), 1.65 μL nucleotide mix (Life Sciences NECN-24; 6X), 0.2 μL of each DNA primer (IDT, 12.5 μM) and 2.3 μL RNA input were assembled at room temperature. After being incubated at 65 °C for 2 min and a 5 min incubation at 41 °C, 2.5 μL of enzyme mix (Life Sciences, NEC-1-24) was added to the reaction. The reaction took place at 41 °C for 80 min, then 10 μL of amplification product was applied to the FARSIGHT reactions with the FARSIGHT sensor and cognate fluorogen (1 μL Corn/2 μM DFHO, 1 μL Red Broccoli FARSIGHT/4 μM OBI, or 1 μL Broccoli/4 μM DFHBI-1T).

### Simultaneous detection of mutations at two sites from a NASBA product

The 10X fluorogenic dye mix in this reaction is consisted of 40 μM DFHBI-1T (Lucerna, 410), 40 μM OBI (Lucerna, 610), 40 mM HEPES (Gibco™, 15630080), pH 7.4, 100 mM KCl (Invitrogen™, AM9640G), and 5 mM MgCl_2_ (Invitrogen™, AM9530G).

The reaction for simultaneously detecting two point mutations from NASBA product was prepared for according to the following recipe: 3.5 μL 10X fluorogenic dye mix, 1.0 µL transcribed Broccoli FARSIGHT, 1.0 µL transcribed Red Broccoli FARSIGHT, 10 µL NASBA product, 19.5 µL DNase/RNase-Free Distilled Water (Invitrogen™, 10977023). The reaction was then incubated in a plate in a temperature-controlled plate reader (Agilent, NEO2MALPHAT) with the Broccoli/DFHBI-1T (Excitation: 472/20, Emission: 507/20) and Red Broccoli/OBI (Excitation: 541/20, Emission: 590/20) fluorescence read out taken in real time at a 37°C constant temperature.

### Clinical virus sample collection and processing

Clinical saliva samples were obtained from the Biodesign Institute Clinical Testing Laboratory (Arizona State University). The specimens were collected for SARS-CoV-2 diagnostic purposes with the consent of the patients and provided as de-identified clinical remnants under the oversight of the Arizona State University Institutional Review Board (IRB ID: STUDY00011737). The viral RNA was extracted using PureLink^TM^ Viral RNA/DNA Mini Kit (Thermo Fisher, 12280050) according to the manufacturer’s instructions. RNA was eluted with 50 µl H_2_O and stored at −80°C before use.

### Reference RT-qPCR reactions

Parallel RT-qPCR detection was implemented using the Luna® SARS-CoV-2 RT-qPCR Multiplex Assay Kit Detection (NEB, E3019). The detection procedure was conducted according to the protocol provided by the kit in 7900HT Fast Real-Time PCR System with 384-Well Block Module (Thermo Fisher, 4329001). Time to threshold was calculated using single threshold analysis mode.

### SARS-CoV-2 sequencing

Whole genome sequencing was performed on saliva specimens RNA extracts as previously described^84^. In brief, library preparation was performed using COVIDSeq Test (Illumina, 20043675) and sequenced on the Illumina NextSeq 2000 instrument using 2 x 109 paired end reads. Sequencing reads were trimmed and quality-filtered to assemble high quality genomes. Sequence quality was validated using VADR (version 1.4)^85^, and lineage calling was performed using Pangolin^86^. All genome sequences were deposited to the GISAID repository.

### Statistical methods

Plate reader dataset analysis and illustrations were performed using MATLAB, SnapGene Viewer, GraphPad Prism, Microsoft Excel, and Adobe Illustrator. Averages of plate reader measurements taken for FARSIGHT reactions were conducted using the arithmetic mean of each technical replicate. Error bars show the standard deviation from three biological technical replicates (*n* = 3) for the fluorescent measurements. Two-tailed Student’s t test calculations were conducted to determine statistical significance in GraphPad Prism. The 95^th^ percentile calculation to assess the diagnostic sensitivity, specificity and accuracy of the FARSIGHT assay were produced using Microsoft Excel.

## Acknowledgments

We thank the Biodesign Institute clinical testing laboratory (Arizona State University) for providing COVID-19-relevant patient samples, and Alexis Thomas, Winston Matthews, Remington Fitch, LaRinda Holland and Matthew Smith for assistance with sequencing. This work was developed with funding from the Defense Advanced Research Projects Agency (DARPA), Contract No. N66001-23-2-4042; a National Institutes of Health (NIH) Director’s New Innovator Award (1DP2GM126892), U01 award (1U01AI148319-01), and R01 award (1R01EB031893); a National Science Foundation (NSF) RAPID award (2029532); Arizona Biomedical Research Centre funds (ADHS16-162400, CTR051763); Canadian Food Inspection Agency funds (39903-200137) and Boston University startup funds to A.A.G. The views, opinions and/or findings expressed are those of the authors and should not be interpreted as representing the official views or policies of the Department of Defense or the U.S. Government. The content is solely the responsibility of the authors and does not necessarily represent the official views of the National Institutes of Health.

## Data availability

The main data supporting the results in this study are available within the main text and the Supplementary Information. The datasets generated during and/or analyzed during the current study are available from the corresponding authors on reasonable request.

## Author contributions

Z.Y. and A.A.G. conceived and designed the studies and experiments. Z.Y. performed most of the wet experiments. Y.L. contributed to RT-qPCR on SARS-CoV-2 clinical samples, A.E. contributed to the screening of optimal NASBA primers, and K.W. contributed to selecting hallmark mutations of Omicron variants. Z.M.T. and V.M. acquired the SARS-CoV-2 clinical samples and E.S.L. performed genomic sequencing of the clinical samples. F.H. designed the biochemical modeling simulation. A.A.G. and F.H. supervised the research. Z.Y. performed the analysis of data. Z.Y., F.H. and A.A.G. wrote and edited the manuscript.

