## Supplementary Information for "Programmable Fluorescent Aptamer-Based RNA Switches for Rapid Identification of Point Mutations"

Supplementary Fig. 13 l Time-course measurements of fluorescence from dual FARSIGHTs following NASBA with 12 positive Omicron BA.5 clinical saliva samples, Related to Fig. 6... 22

Supplementary Fig. 14 l Fluorescence measurements of the dual FARSIGHT assay following NASBA with 11 negative clinical patient saliva samples, Related to Fig. 6............................ 23

*Supplementary References*.................................................................................................... *24*

### Supplementary Note 1: Biochemical modeling of FARSIGHT performance

To better design and understand FARSIGHT performance, we performed biochemical modeling of their behavior. We did kinetic simulations to study the transition of the two strand-displacement reactions as bellow:

$$OFF\ state \xrightarrow{k_{on}} INT\ state$$
$$INT\ state + Fluorogen \xrightarrow{k_{asso}} ON\ state$$

The ordinary differential equations for the reactions are the following:

$$\frac{d(RNA-OFF)}{dt} = -k_{on}[RNA-OFF] + k_{off}[RNA-INT]$$
$$\frac{d[RNA-INT]}{dt} = k_{on}[RNA-INT] - k_{off}[RNA-INT] - k_{asso}[RNA-INT][Fluorogen] + k_{asso-off}[RNA-ON]$$
$$\frac{d[Fluorogen]}{dt} = -k_{asso}[RNA-INT][Fluorogen] + k_{asso-off}[RNA-ON]$$
$$\frac{d[RNA-ON]}{dt} = k_{asso}[RNA-INT][Fluorogen] - k_{asso-off}[RNA-ON]$$

During the first step of the strand displacement reactions, the relationship between the $k_{on}$ and $k_{off}$ is:

$$\frac{k_{on}}{k_{off}} = e^{\frac{-\Delta G_{OFF-INT}}{RT}}$$

Where $\Delta G_{OFF\text{-}INT}$ is the reaction equilibrium energy of the first strand-displacement reaction with mutant target:

$$\Delta G_{OFF\text{-}INT} = \Delta G_{INT} - \Delta G_{OFF}$$

However, in the case of wild-type (WT) target binding, with a mismatch present in the intermediate because of a single-nucleotide mutation, there will be a $\Delta G_{bulge}$ energy penalty of ~4 kcal/mol for a single-nucleotide bulge[2,3]. The equilibrium energy in the case of mismatch is:

$$\Delta G_{OFF\text{-}INT\text{-}WT} = \Delta G_{OFF\text{-}INT} + \Delta G_{bulge}$$

During the second step of the strand displacement reactions and fluorogen binding, the relationship between the $k_{asso}$ and $k_{asso\text{-}off}$ is:

$$\frac{k_{asso}}{k_{asso-off}} = e^{\frac{-\Delta G_{asso}}{RT}}$$

Where $\Delta G_{asso}$ is the reaction equilibrium energy of the second strand-displacement reaction and fluorogen binding. As the second step involves strand displacement and fluorogen binding, $\Delta G_{asso}$ can be calculated by the addition of equilibrium energy without fluorogen ($\Delta G_{INT\text{-}ON}$) and the binding energy of the fluorogen to the RNA aptamer ($\Delta G_{fluorogen}$):

$$\Delta G_{INT\text{-}ON} = \Delta G_{ON} - \Delta G_{INT}$$
$$\Delta G_{asso} = \Delta G_{INT\text{-}ON} + \Delta G_{fluorogen}$$

when the reaction is completed, the differentiation factor (Df) is calculated by the following equation:

$$Df = \frac{[\mathrm{RNA} - \mathrm{ON_{mut}}]_{end} + Background}{[RNA - ON_{wt}]_{end} + Background}$$

Where $[\mathrm{RNA\text{-}ON_{mut}}]_{end}$ and $[\mathrm{RNA\text{-}ON_{wt}}]_{end}$ are the final ON state of the FARSIGHT when bound with the mutant and wild-type RNA target, respectively. As there is a detection limit for the instrument to read the fluorescent signal from the final ON state, a background is added, which is 5% of the original FARSIGHT concentration.

The rate constants for the two steps are estimated to be $10^6$ $s^{-1}$ and $10^3$ $M^{-1}s^{-1}$, respectively. To solve the ordinary different equation set, MATLAB ode23s solver was used with the following options for the tolerance values:

```
options = odeset('RelTol', 1e-4, 'AbsTol', 1e-30);
```

### Supplementary Note 2: FARSIGHT in silico design and FARSIGHT/target interaction energy calculation

The competitive reactions occurring between ON state ("$\Delta G_{ON}$"), intermediate state ("$\Delta G_{INT}$"), and OFF state ("$\Delta G_{OFF}$") of FARSIGHT systems upon binding to the target are critical for discrimination of single-nucleotide mutations. The complex subsequently can transition from OFF state to INT state and to ON state, and these processes are designed to be reversible. The equilibrium between these three states can be represented by the following chemical reaction.

$$OFF\ state \rightarrow INT\ state\ \rightarrow ON\ State$$
$$K_{eq} = \frac{State\ 2}{State\ 1} = e^{\frac{-\Delta G}{RT}}$$

First, to maximize the chances of obtaining the best design, the design algorithm takes in the target mutation and WT sequence to be distinguished, the core sequence of the output aptamer (as listed below) and fluorogen to generate a library of FARSIGHT designs in silico.

| | |
|---|---|
| Broccoli aptamer (rotated configuration) | GUCGAGUAGAGUGUGGNNNNNNNNNNNNNNNNNNNNCGGUCGGGUCC<br>(..((.(.((...(.(((((((((....))))))))).))).).))) |
| Broccoli aptamer (standard configuration) | CGGUCGGGUCCNNNNNNNNNNNNNNNNNNNNGUCGAGUAGAGUGUGG<br>(.(((.(.(((((((((((....)))))))))..)).).))...).) |
| Red Broccoli aptamer (standard configuration) | CGGUCGGGUCCNNNNNNNNNNNNNNNNNNNNGUUGAGUAGUGUGUGG<br>(.(...(.(((((((((((....)))))))))..)).).......).) |
| Red Broccoli aptamer (rotated configuration) | GUUGAGUAGUGUGUGGNNNNNNNNNNNNNNNNNNNNCGGUCGGGUCC<br>(..((.(.......(.(((((((((....))))))))).)...).))) |
| Corn aptamer | CGAGGAAGGAGGUCUGAGGAGGUCACUG<br>(.....((..(..((....))..).))) |

Then, the designs are sorted by predicted performance using a scoring function considering (i) overall ensemble defect of the design secondary structure, (ii) the reaction energy, and (iii) energy contribution from ligand binding. Take DFHBI-1T[1] for example:

$$K_d = 560\ nM$$
$$\Delta G_{Fluorogen} = -RT\ln(K_d) = -8.8723\ kcal/mol$$

In a desired reaction where the FARSIGHT binds to the target with the correct sequence, Gibbs free energy transition $\Delta G_{OFF\text{-}INT}$ is expected to be slightly negative, yielding equilibrium shifting to the intermediate state and eventual progression to the ON state. However, a point mutation in the target is assumed to cause a very positive energy penalty (~4 kcal/mol)[2,3], making equilibrium shift toward the OFF state.

Finally, the top-ranked FARSIGHT designs with the optimal scores were chosen for experimental validation. All prospective FARSIGHTs designs for a given combination of forward and reverse toehold lengths shared identical secondary structures but varied in sequence outside of the conserved sequence regions of the aptamer.

### Supplementary Note 3: Estimate of FARSIGHT costs

The costs attributed to the reaction readout for FARSIGHTs comprise: (1) the dsDNA transcription template, (2) the *in vitro* transcription reaction to produce FARSIGHT RNA, (3) the fluorogen for binding to activate FARSIGHTs. NASBA reactions are required for amplification of target RNA. The costs for a 20-$\mu$L FARSIGHT reaction are calculated in the following tables.

(1) DNA template cost

| FARSIGHT | DNA template sequence | Length (bp) | Yield (nmoles) | Cost/ Base (dsDNA) | Total Cost of dsDNA | Concentration Used for Transcription ($\mu$M) | Transcription Volume per Assay ($\mu$L) | Nanomoles Used per Assay | **Cost Per Assay** |
|---|---|---|---|---|---|---|---|---|---|
| rot_broc_S_gene_N501Y_AS_SNIPA_M5_X4x5_Y4x3_R01 | GCGCTAATACGACTCACTATAGGGACTTTCCTTTACAATCATATGCTTCTAAACACACTTATGGTGTTGGTTACCACGTAAAAAATAACAAAACACTAAATACGTGGTAACCAACACCATATCGAGTAGAGTGTGGGCTCAGATTCGTCTGAGACGGTCGGGTCTATGGTGTTGGTTTATAATAACCAACAC | 192 | 20 | $3.86 | $741.12 | 0.2 | 1.0 | 0.0002 | **$0.0074** |
| std_red_broc_S_gene_Y505H_AS_SNIPA_a3_b3_c07_d5_e3_f3_M3_dg2_R01 | GCGCTAATACGACTCACTATAGGGTTTCCTTTACGATCATATAGTTTAGAACACTCTTATGGTGTTGGTCACCAACTGGTCAAATACATGCAATTAACTCCAGTTGGTGACCAACACCATCGGTCGGGTCCAATCTTGTGTGGACAAGGTTGTTGAGTAGTGTGTGGATGGTGTTGGTCACCGATATGGGTGACCAACAC | 200 | 20 | $3.86 | $772.00 | 0.2 | 1.0 | 0.0002 | **$0.0077** |

Note: DNA costs are from IDT using the ultramer 20-nmole synthesis scale to generate a double-stranded DNA template for transcription.

(2) *In vitro* transcription cost

| Transcript | Kit Cost | Kit Reaction Volume ($\mu$L) | Cost per $\mu$L of Transcription Reaction | Transcription Volume per Assay ($\mu$L) | **FARSIGHT Transcription Cost per Assay** |
|---|---|---|---|---|---|
| Red Broccoli FARSIGHT | $512 | 1000 | $0.51 | 1.0 | **$0.51** |
| Broccoli FARSIGHT | $512 | 1000 | $0.51 | 1.0 | **$0.51** |

Note: Transcription cost was determined using the Biosearch Technologies ASF3507 kit. Transcription costs will be much lower using a homemade *in vitro* transcription reaction.

(3) Fluorogen cost

| Fluorogen | Molecular Weight (g/mol) | Price | Mass from Vendor (mg) | Moles from Vendor | Concentration in Assay ($\mu$M) | Assay Volume ($\mu$L) | Moles per Assay | **Cost per Assay** |
|---|---|---|---|---|---|---|---|---|
| OBI | 397.34 | $199.99 | 1 | 2.51674E-06 | 4 | 20 | 8E-11 | **$0.0064** |
| DFHBI-1T | 320.21 | $299.99 | 10 | 3.12295E-05 | 4 | 20 | 8E-11 | **$0.0008** |

Note: Fluorogen costs are from Lucerna.

(4) NASBA primer cost

| FARSIGHT | DNA template sequence | Length (bp) | Yield (nmoles) | Cost/ Base (dsDNA) | Total Cost of dsDNA | Concentration Used for Transcription ($\mu$M) | Transcription Volume per Assay ($\mu$L) | Nanomoles Used per Assay | **Cost Per Assay** |
|---|---|---|---|---|---|---|---|---|---|
| NASBA_Omicron_S_gene_Q493R_to_Y505H_AS_0061_fwd | AATTCTAATACGACTCACTATAGGGAGAAGGCACAAACAGTTGCTGGTGCA | 51 | 25 | $0.43 | $21.93 | 0.25 | 10.0 | 0.0002 | **$0.0022** |
| NASBA_Omicron_S_gene_Q493R_to_Y505H_AS_0061_rev | AATCTATCAGGCCGGTAACA | 20 | 25 | $0.43 | $8.60 | 0.25 | 10.0 | 0.0002 | **$0.0009** |

Note: DNA costs are from IDT using the DNA oligo 25-nmole synthesis scale for NASBA amplification.

(5) NASBA reagent cost

| NASBA reagents | Kit Cost | Kit Reaction Volume ($\mu$L) | Cost per $\mu$L of NASBA Reaction | NASBA reagent Volume per Assay ($\mu$L) | NASBA primer cost | **NASBA Cost per Assay** |
|---|---|---|---|---|---|---|
| NASBA Liquid Kit | $386.65 | 120 | $3.22 | 7.5 | 0.0031 | **$24.1687** |

Note: NASBA reagent costs are from Life Sciences.

Combined Readout Costs for One- and Two-Channel FARSIGHT Assays (commercially available kits)

| | (1) DNA Template | (2) *In vitro* Transcription | (3) Fluorogen | **Total Cost** | Assay Type |
|---|---|---|---|---|---|
| Red Broccoli FARSIGHT | $0.0074 | $0.51 | $0.0064 | **$0.5238** | One-Channel |
| Broccoli FARSIGHT | $0.0077 | $0.51 | $0.0008 | **$0.5185** | One-Channel |
| Red Broccoli and Broccoli FARSIGHTs | $0.0151 | $1.02 | $0.0072 | **$1.0423** | Two-Channel |

The readout costs for the FARSIGHT reactions are thus ~$0.52 per assay per FARSIGHT used. 98% of this cost is due to transcription, which could be reduced using homemade transcription reactions.

Combined Costs for NASBA-coupled One- and Two-Channel FARSIGHT Assays (commercially available kits)

| | (1) NASBA | (2) Readout | **Total Cost** | Assay Type |
|---|---|---|---|---|
| Red Broccoli FARSIGHT | $24.1687 | $0.5238 | **$24.6925** | One-Channel |
| Broccoli FARSIGHT | $24.1687 | $0.5185 | **$24.6872** | One-Channel |
| Red Broccoli and Broccoli FARSIGHTs | $24.1687 | $1.0423 | **$25.2110** | Two-Channel |

The costs for the FARSIGHT reactions using commercial transcription and NASBA kits are thus ~$24.69 per assay per FARSIGHT used. 98% of this cost is due to NASBA, which could be reduced using homemade NASBA reactions.

(6) Homemade transcription kit cost

| Transcription Reagents | Reagent Cost | Reagent quantity | Quantity used per 20 $\mu$L transcription | Cost per 20 $\mu$L transcription | Cost per $\mu$L of Transcription Reaction | Transcription Volume per Assay ($\mu$L) | **FARSIGHT Transcription Cost per Assay** |
|---|---|---|---|---|---|---|---|
| T7 RNA Polymerase (High Concentration)/reaction Buffer, NEB | $610.00 | 50,000 units | 40 units | $0.4880 | $0.02440 | 1.0 | **$0.02440** |
| RNase Inhibitor, Murine, NEB | $322.00 | 15,000 units | 20 units | $0.4293 | $0.02147 | 1.0 | **$0.02147** |
| Ribonucleotide Solution Mix, NEB | $333.00 | 50 $\mu$mol | 0.5 mM | $0.3330 | $0.01665 | 1.0 | **$0.01665** |
| DTT (dithiothreitol), Thermo Fisher | $415.65 | 0.16 mol | 5 mM | $0.0003 | $0.00001 | 1.0 | **$0.00001** |
| Total | | | | | $0.06253 | 1.0 | **$0.06253** |

(7) Homemade NASBA kit cost

| NASBA Reagents | Reagent Cost | Reagent quantity | Quantity used per 25 $\mu$L NASBA reaction | Cost per 25 $\mu$L NASBA reaction | NASBA Volume per Assay ($\mu$L) | **Cost per Assay** |
|---|---|---|---|---|---|---|
| AMV Reverse Transcriptase, NEB | $352.00 | 1,000 units | 8 units | $2.8160 | 10.00 | **$1.1264** |
| T7 RNA Polymerase (High Concentration), NEB | $610.00 | 50,000 units | 40 units | $0.4880 | 10.00 | **$0.1952** |
| Thermostable RNase H, NEB | $163.00 | 250 units | 0.2 units | $0.1304 | 10.00 | **$0.0522** |
| Rnase Inhibitor, Murine, NEB | $322.00 | 15,000 units | 12.5 units | $0.2683 | 10.00 | **$0.1073** |
| dNTP mixture, NEB | $288.00 | 40 $\mu$mol | 1 mM | $0.1800 | 10.00 | **$0.0720** |
| Ribonucleotide Solution Mix, NEB | $333.00 | 50 $\mu$mol | 2 mM | $0.3330 | 10.00 | **$0.1332** |
| Tris-HCl (pH 8.5), Thermo Fisher | $116.00 | 0.5 mol | 40 mM | $0.0002 | 10.00 | **$0.0001** |
| KCl, Thermo Fisher | $52.65 | 0.2 mol | 50 mM | $0.0003 | 10.00 | **$0.0001** |
| MgCl2, Thermo Fisher | $53.65 | 0.1 mol | 12 mM | $0.0002 | 10.00 | **$0.0001** |
| DTT (dithiothreitol), Thermo Fisher | $415.65 | 0.16 mol | 10 mM | $0.0006 | 10.00 | **$0.0003** |
| DMSO (Dimethyl sulfoxide), Thermo Fisher | $110.00 | 100% x 1 L | 0.15 | $0.0004 | 10.00 | **$0.0002** |
| BSA, Thermo Fisher | $292.65 | 150 mg | 100 µg/ml | $0.0049 | 10.00 | **$0.0020** |
| Total | | | | | | **$1.6890** |

Note: We refer to Sooknanan et al., Molecular methods for virus detection. 261-285 (1995) for homemade NASBA assembly[4]. NASBA reagent costs are either NEB or Thermo Fisher.

Combined Costs for NASBA-coupled One- and Two-Channel FARSIGHT Assays (homemade transcription and NASBA reactions)

| | (1) DNA Template | (2) *In vitro* Transcription | (3) Fluorogen | (4) NASBA | **Total Cost** | Assay Type |
|---|---|---|---|---|---|---|
| Red Broccoli FARSIGHT | $0.0074 | $0.06253 | $0.0064 | $1.6890 | **$1.76533** | One-Channel |
| Broccoli FARSIGHT | $0.0077 | $0.06253 | $0.0008 | $1.6890 | **$1.76003** | One-Channel |
| Red Broccoli and Broccoli FARSIGHTs | $0.0151 | $0.12506 | $0.0072 | $1.6890 | **$1.83636** | Two-Channel |

**Supplementary Figures**

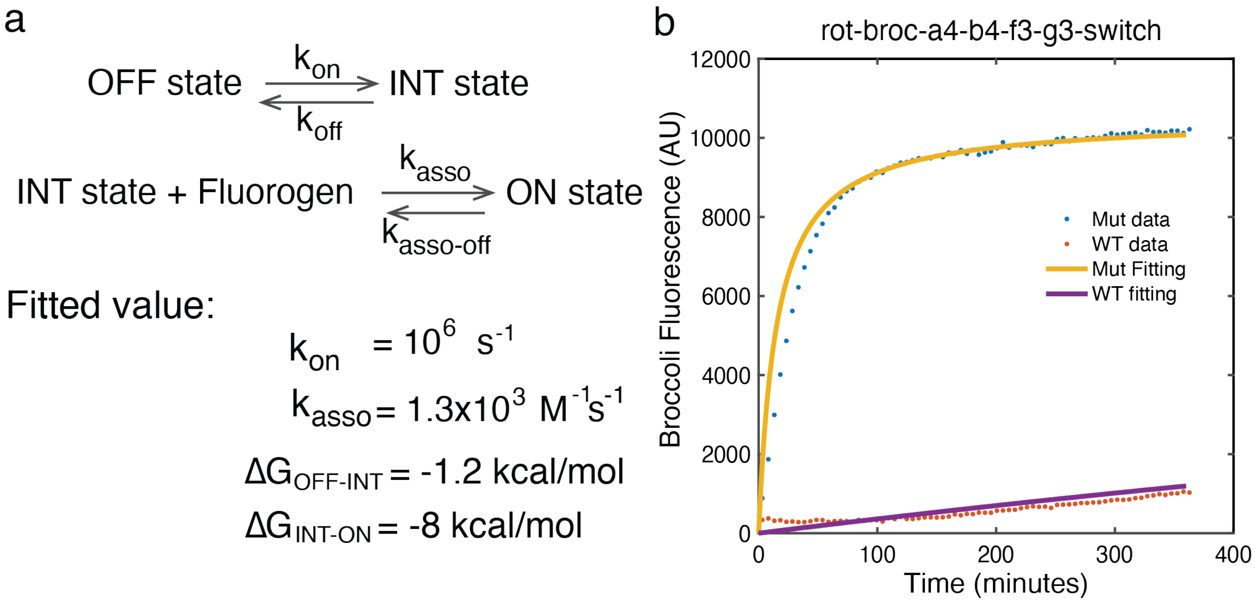

**Supplementary Fig. 1 I The two-step kinetic modeling of FARSIGHT performance and fitting to the experimental data.**

**a**, The chemical equation for the two-step reactions and fitted parameter values for the design in panel b.

**b**, The experimental data and fitted curve shown for a FARSIGHT with Broccoli output.

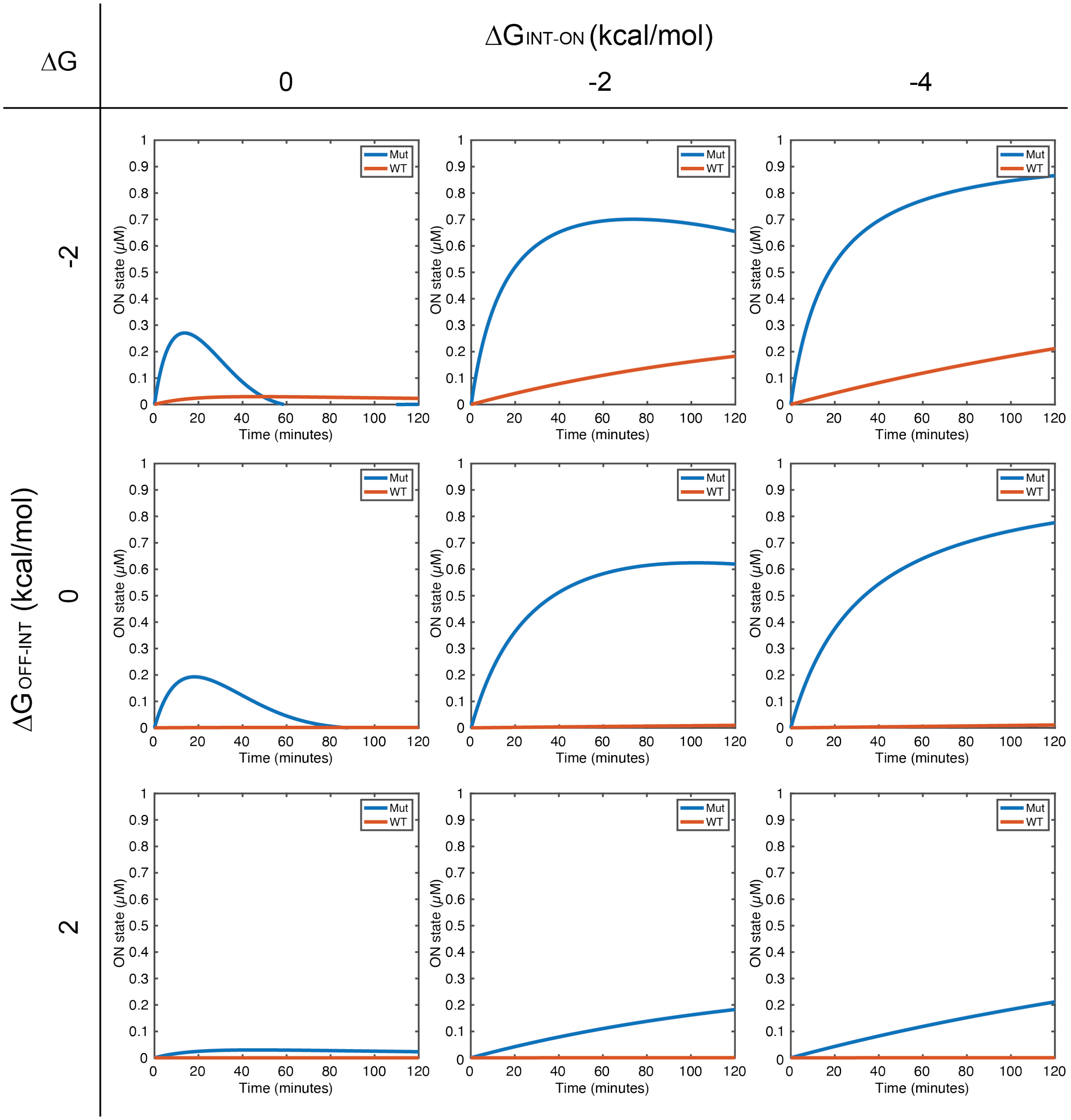

**Supplementary Fig. 2 | Simulated kinetic curves for different combinations of equilibrium energies for FARSIGHT two-step reactions.**

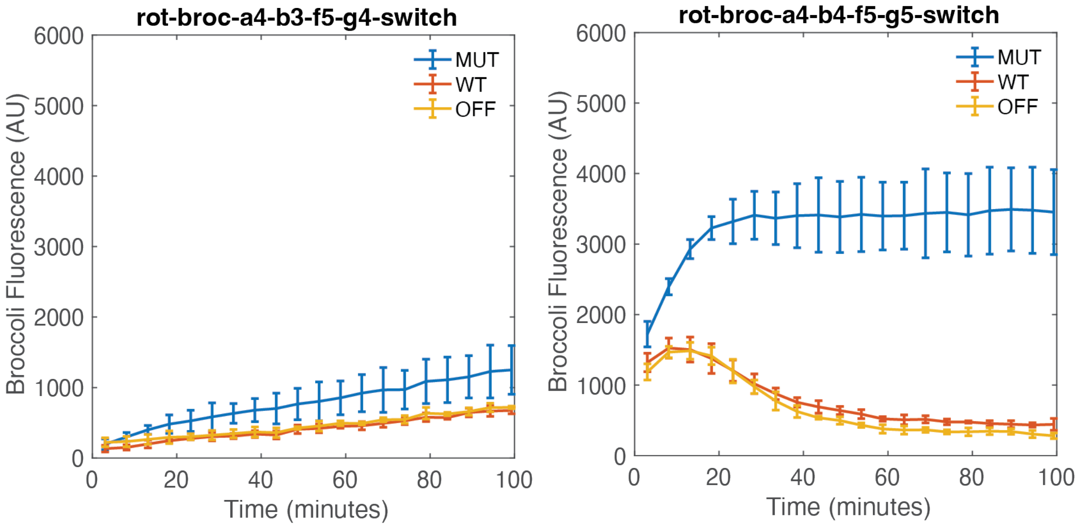

**Supplementary Fig. 3 | Two sample FARSIGHT designs that have the same equilibrium energy based on thermodynamic models but substantially different performance in experiments.**

a

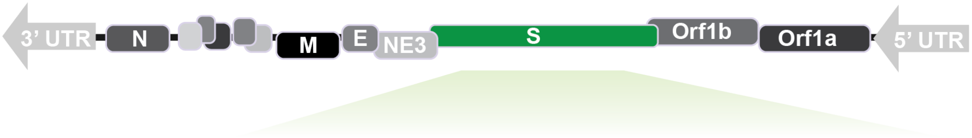

| SARS-CoV-2 Main Variants | | Δ69/70 | K417N | K417T | V445P | E484K | E484Q | F490S | Q493R | G496S | Q498R | N501Y | Y505H | P681R | P681H | N969K | V1176F |
|---|---|---|---|---|---|---|---|---|---|---|---|---|---|---|---|---|---|
| B.1.1.7 (Alpha) | | + | | | | | | | | | | + | | | + | | |
| B.1.351 (Beta) | | | + | | | + | | | | | | + | | | | | |
| P.1 (Gamma) | | | | + | | + | | | | | | + | | | | | + |
| B.1.617 | B.1.617.1 (Kappa) | | | | | | + | | | | | | | + | | | |
| | B.1.617.2 (Delta) | | | | | | | | | | | | | + | | | |
| B.1.1.529 (Omicron) | XBB | | + | | + | | | + | | | + | + | + | | + | + | |
| | BA.1 | + | + | | | | | | + | + | + | + | + | | + | + | |
| | BA.2 | | + | | | | | | + | | + | + | + | | + | + | |
| | General Omicron | | + | | | | | | | | + | + | + | | + | + | |

b

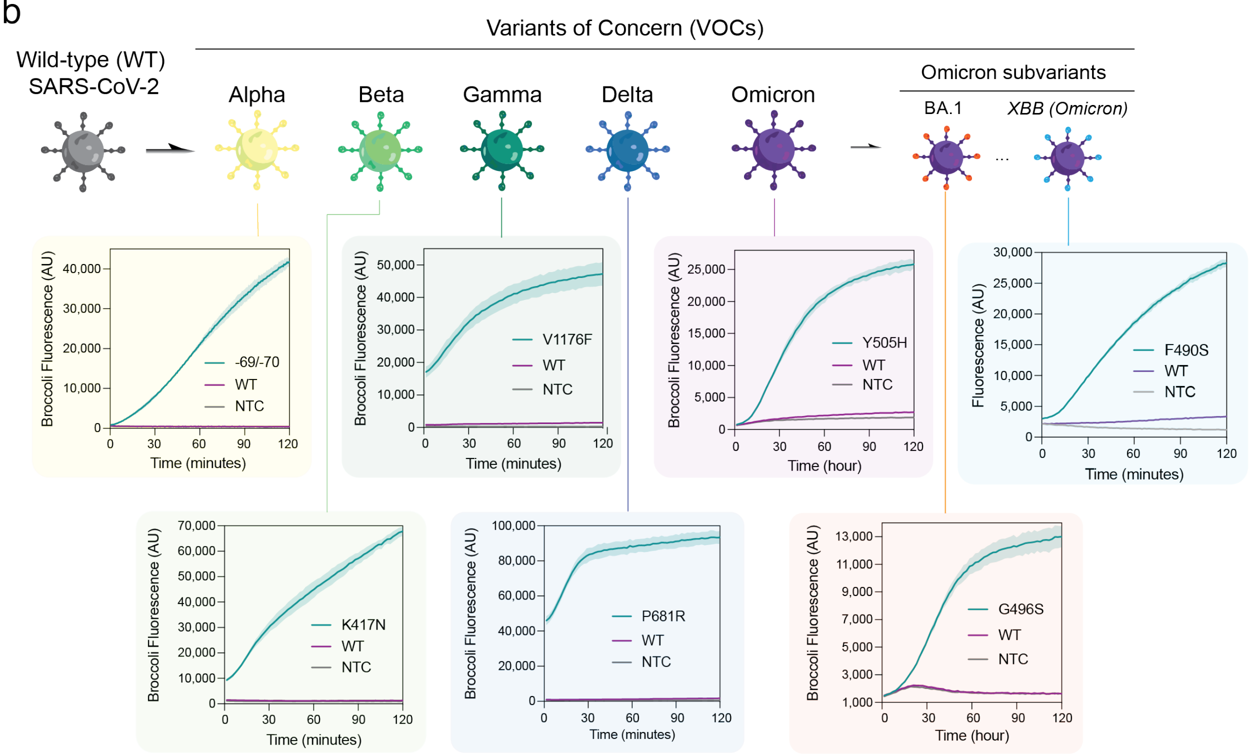

**Supplementary Fig. 4 I FARSIGHT discriminates hallmark mutations in SARS-CoV-2 to genotype variants of interest.**

**a**, Mutation distribution for different important SARS-COV-2 variants including Alpha, Beta, Gamma, Kappa, Delta, Omicron, and Omicron subvariants. Hallmark mutation set was used to characterize specific SARS-CoV-2 variants (shown in green).

**b**, Time-course measurements of fluorescence from Broccoli FARSIGHT systems for recognition of different SARS-CoV-2 variants. Shaded regions denote mean ± s.d. with *n*=3 technical replicates.

See **Supplementary Table 4**.

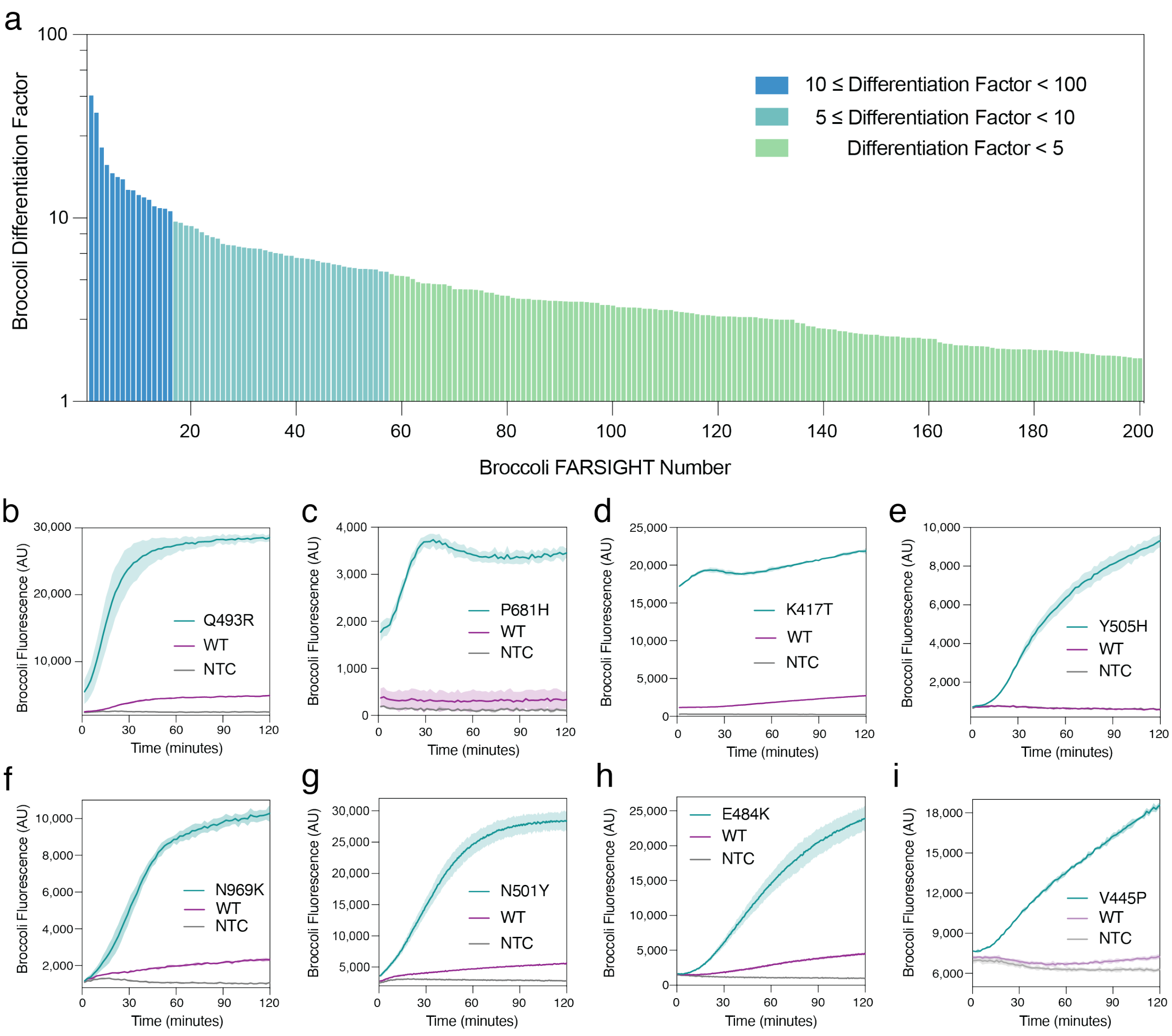

**Supplementary Fig. 5 I Characterization of FARSIGHTs for SARS-CoV-2 hallmark mutation detection.**

**a**, Differentiation factors of 180 Broccoli FARSIGHTs that discriminate mutant targets from the wild-type sequence.

**b-i**, Fluorescence measurements as a function of time for representative Broccoli FARSIGHTs featuring RNA with target mutations, the wild-type sequence, and those without the SARS-CoV-2 target RNA. Measurements were taken after 2 hours of FARSIGHT detection. Shaded regions denote mean ± s.d. with *n*=3 technical replicates.

See **Supplementary Table 3**.

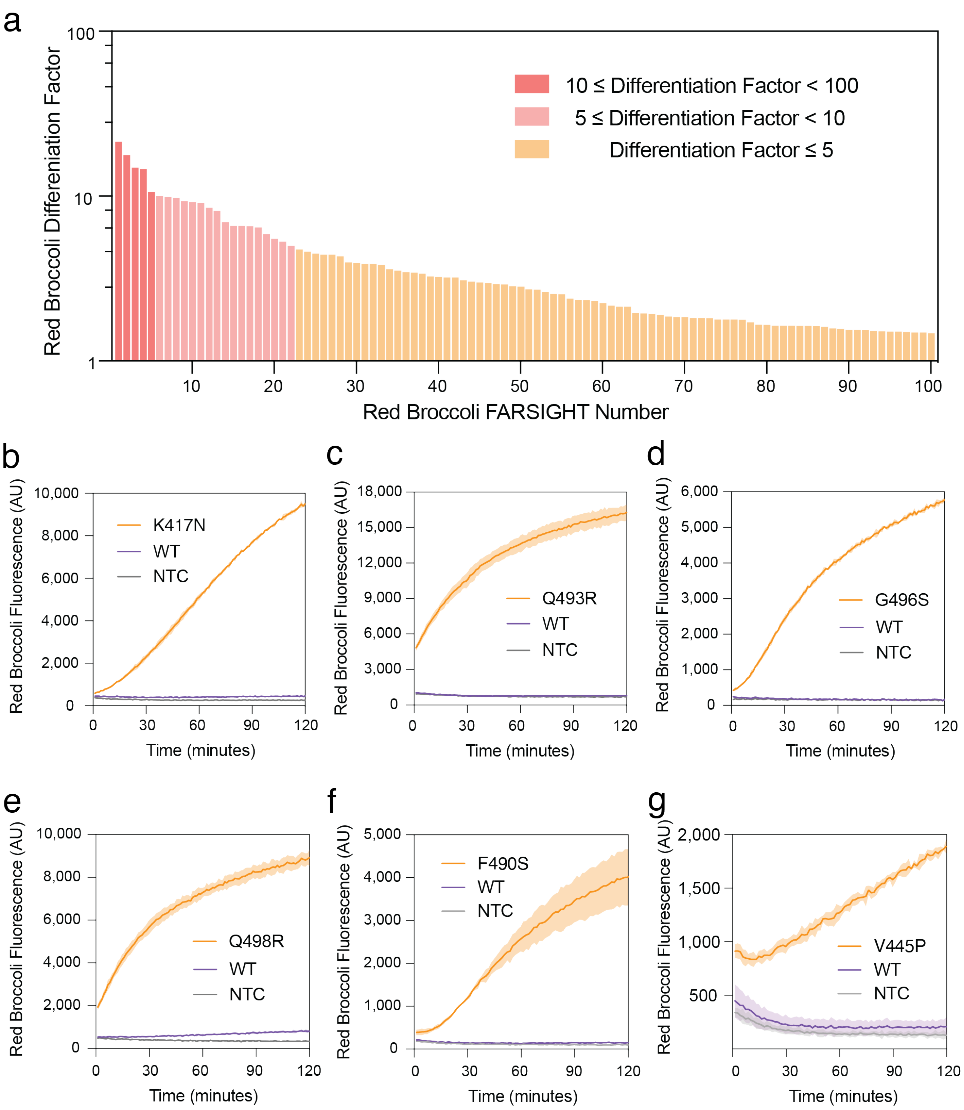

**Supplementary Fig. 6 I Screening of Red Broccoli FARSIGHTs for SARS-CoV-2 hallmark mutation detection.**

**a**, Differentiation factor of 80 Red Broccoli FARSIGHTs that discriminate mutant targets from the wild-type sequence.

**b-g**, Time-course measurements of fluorescence from representative Red Broccoli FARSIGHTs with target mutation-carrying RNA and wild-type sequence and without the SARS-CoV-2 target RNA. Measurements were taken after 2 hours of FARSIGHT detection. Shaded regions denote mean ± s.d. with *n*=3 technical replicates.

See **Supplementary Table 5**.

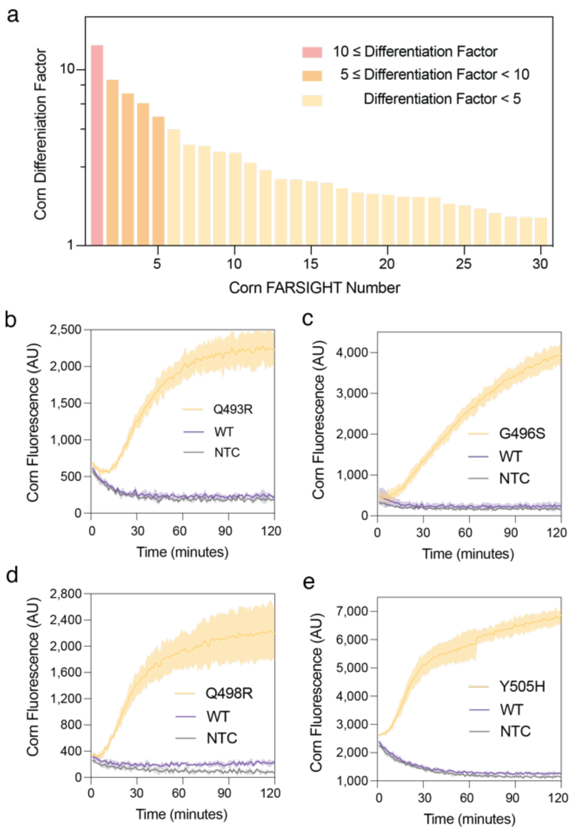

**Supplementary Fig. 7 I In vitro screening of Corn FARSIGHTs for SARS-CoV-2 hallmark mutation detection.**

**a**, Differentiation factors of Corn FARSIGHTs that discriminate correct mutant targets from the wild-type sequence.

**b-e**, Time-course measurements of Corn FARSIGHTs featuring correct mutant target, wild-type sequence and those without the SARS-CoV-2 target RNA. Measurements were taken after 2 hours of FARSIGHT detection. Shaded regions denote mean ± s.d. with *n*=3 technical replicates.

See **Supplementary Table 6**.

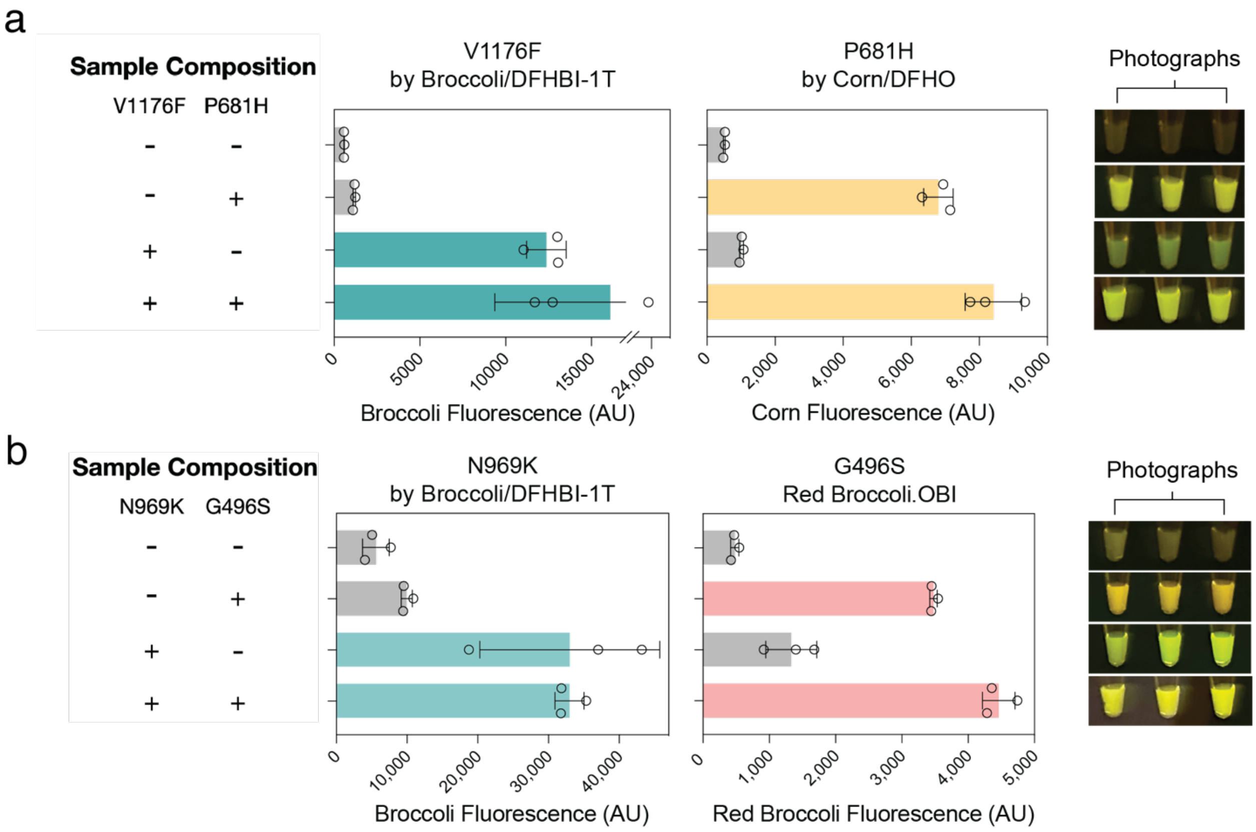

**Supplementary Fig. 8 I Two-channel simultaneous detection of point mutations using spectrally distinct FARSIGHT/fluorogen pairs.**

**a**, Simultaneous detection of V1176F and P681H mutations by Broccoli/DFHBI-1T and Corn/DFHO, respectively. Reactions were measured in triplicate.

**b**, Simultaneous detection of N969K and G496S mutations by Broccoli/DFHBI-1T and Red Broccoli/OBI, respectively. Reactions were measured in triplicate.

See **Supplementary Table 7**.

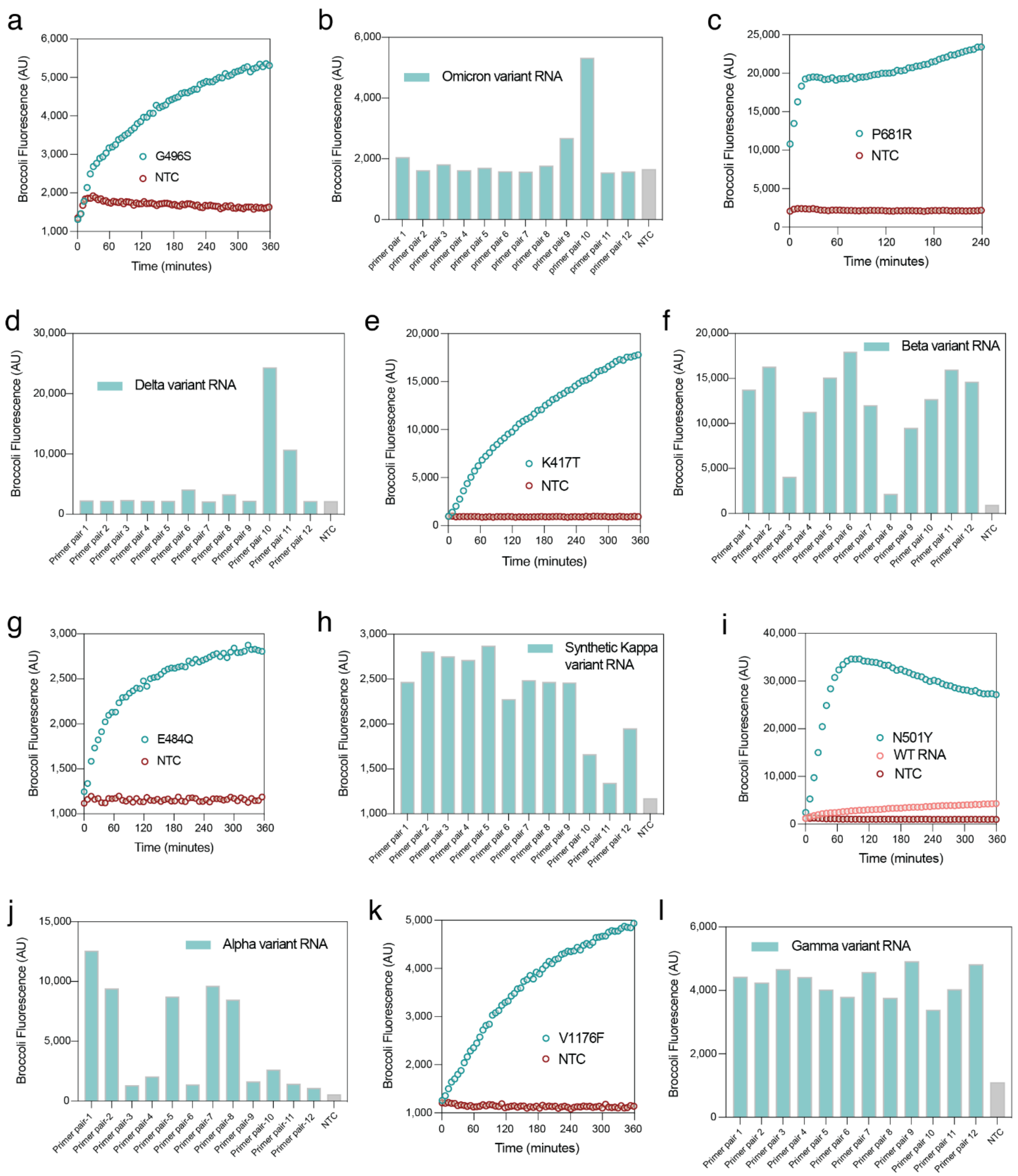

**Supplementary Fig. 9 | Screening results of 12 different NASBA primer pairs from reactions with 200 copies/μL of SARS-CoV-2 variant genomic RNA**

**a**-**i**, Performance evaluation of Broccoli FARSIGHTs targeting G496S (Omicron, **a**-**b**), P681R (Delta, **c**-**d**), K417T (Gamma, **e**-**f**), E484Q (Kappa, **g**-**h**), N501Y (**I**-**j**), and V1176F (Gamma, **k**-**l**) following NASBA with 12 different primers pairs using synthetic SARS-CoV-2 RNA controls (Twist Bioscience).

Primer performance was assessed based on strength of fluorescent response. The mutations that are in close proximity to each other can be amplified using the same primers. **a**, **c**, **e**, **g**, **i** and **k** show time-course measurements of fluorescence using optimal NASBA primer pair. The optimal primer pair was then combined with the FARSIGHT for limit of detection tests.

See **Supplementary Table 8**.

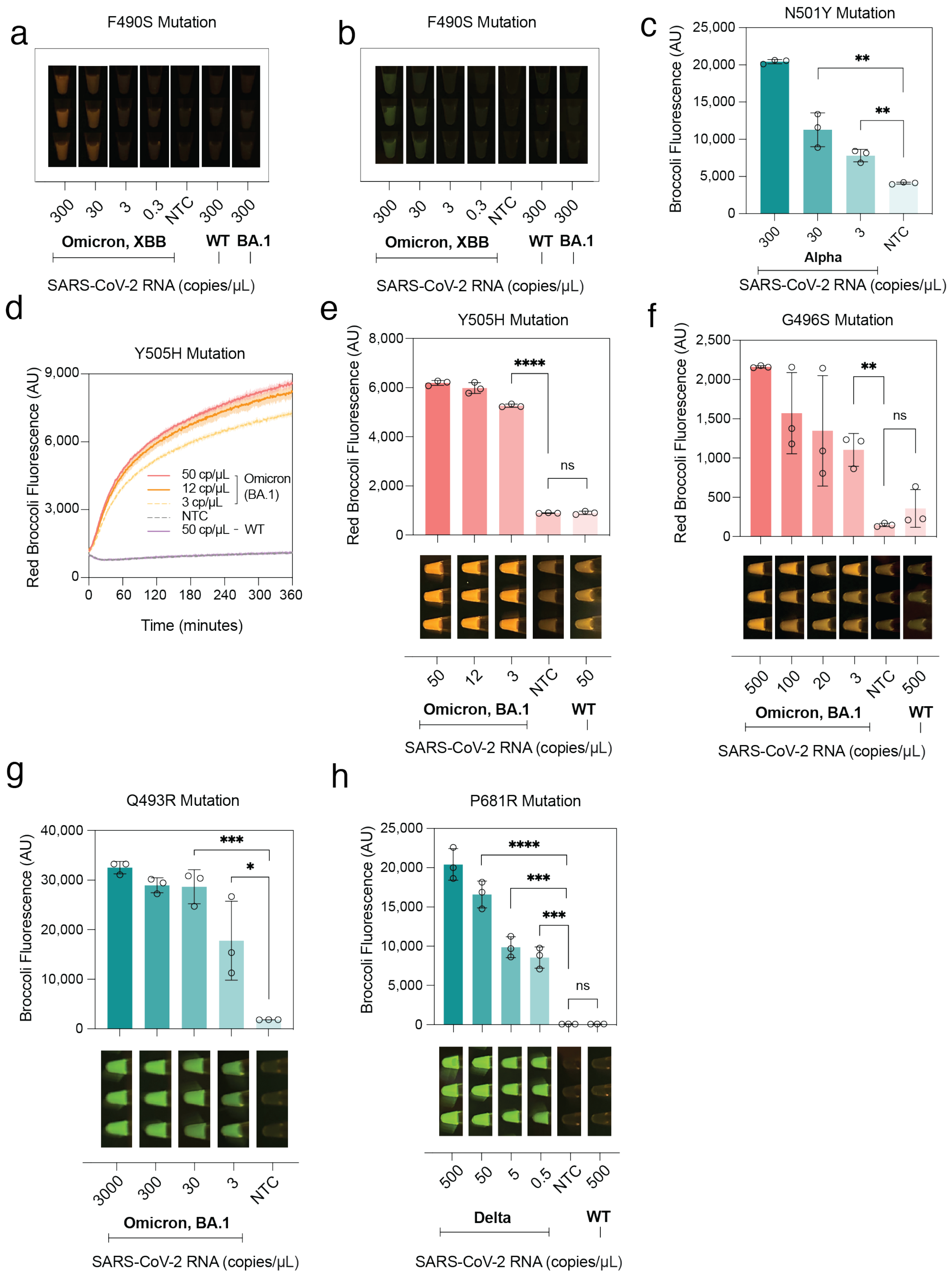
a
F490S Mutation
300 30 3 0.3 NTC 300 300
Omicron, XBB WT BA.1
SARS-CoV-2 RNA (copies/µL)
b
F490S Mutation
300 30 3 0.3 NTC 300 300
Omicron, XBB WT BA.1
SARS-CoV-2 RNA (copies/µL)
c
N501Y Mutation
Broccoli Fluorescence (AU)
25,000
20,000
15,000
10,000
5,000
0
**
**
300 30 3 NTC
Alpha
SARS-CoV-2 RNA (copies/µL)
d
Y505H Mutation
Red Broccoli Fluorescence (AU)
9,000
6,000
3,000
0
50 cp/µL
12 cp/µL
3 cp/µL
NTC
50 cp/µL - WT
Omicron (BA.1)
0 60 120 180 240 300 360
Time (minutes)
e
Y505H Mutation
Red Broccoli Fluorescence (AU)
8,000
6,000
4,000
2,000
0
****
ns
50 12 3 NTC 50
Omicron, BA.1 WT
SARS-CoV-2 RNA (copies/µL)
f
G496S Mutation
Red Broccoli Fluorescence (AU)
2,500
2,000
1,500
1,000
500
0
**
ns
500 100 20 3 NTC 500
Omicron, BA.1 WT
SARS-CoV-2 RNA (copies/µL)
g
Q493R Mutation
Broccoli Fluorescence (AU)
40,000
30,000
20,000
10,000
0
***
*
3000 300 30 3 NTC
Omicron, BA.1
SARS-CoV-2 RNA (copies/µL)
h
P681R Mutation
Broccoli Fluorescence (AU)
25,000
20,000
15,000
10,000
5,000
0
****
***
***
ns
500 50 5 0.5 NTC 500
Delta WT
SARS-CoV-2 RNA (copies/µL)

**Supplementary Fig. 10 I Photograph of fluorescence from FARSIGHT reactions following NASBA of different SARS-CoV-2 RNA variant concentrations.**

**a**-**h**, Limit of detection for the detection of (**a**-**b**) F490S by Broccoli and Red Broccoli FARSIGHTs, (**c**) N501Y by Broccoli Broccoli FARSIGHT, (**d**-**e**) Y505H by Red Broccoli FARSIGHT, (**f**) G496S by Red Broccoli FARSIGHT, (**g**) Q493R by Broccoli FARSIGHT, (**h**) P681R by Broccoli FARSIGHT, and (**g**) N501Y by Broccoli FARSIGHT.

Reactions were measured in triplicate. Shaded regions denote mean ± s.d. with $n$=3 technical replicates. Two-tailed Student t test; ns, $p > 0.05$; **, $p < 0.01$; ***, $p < 0.001$; ****, $p < 0.0001$.

See **Supplementary Table 9**.

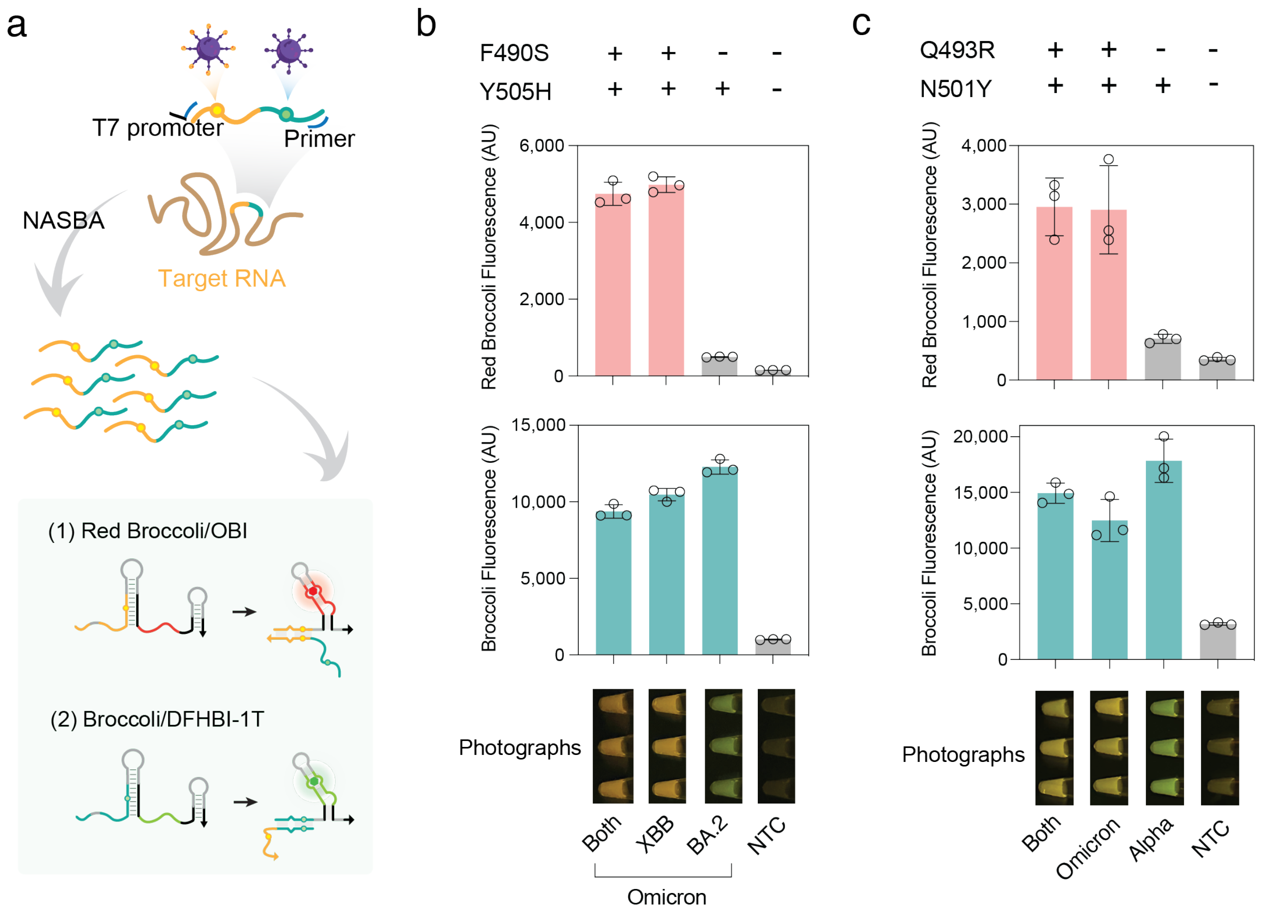

**Supplementary Fig. 11 I Simultaneous identification of SARS-CoV-2 variants from a single NASBA product.**

**a**, Schematic of assay workflow for multiplexed SARS-CoV-2 variant discrimination. A region containing multiple hallmark mutations from the SARS-CoV-2 S gene was chosen for amplification via NASBA. Extracted RNA from patient saliva samples was first amplified by NASBA for 80 minutes at 41°C. The reaction products are then diluted into a second pot containing FARSIGHTs for each mutation and their companion fluorogen. A rapid increase in the fluorescence of Broccoli and/or Red Broccoli FARSIGHTs signifying the presence of respective variants.

**b**, Red Broccoli FARSIGHTs discriminate XBB variant by identifying F490S mutation from general Omicron variants verified by Y505H-targeted Broccoli FARSIGHTs.

**c**, Red Broccoli FARSIGHTs discriminate BA.1 variant by identifying G496S mutation from general SARS-CoV-2 variants verified by N501Y-targeted Broccoli FARSIGHTs.

(*n*=3 technical replicates; bars represent arithmetic mean ± s.d.)

See **Supplementary Table 10**.

a

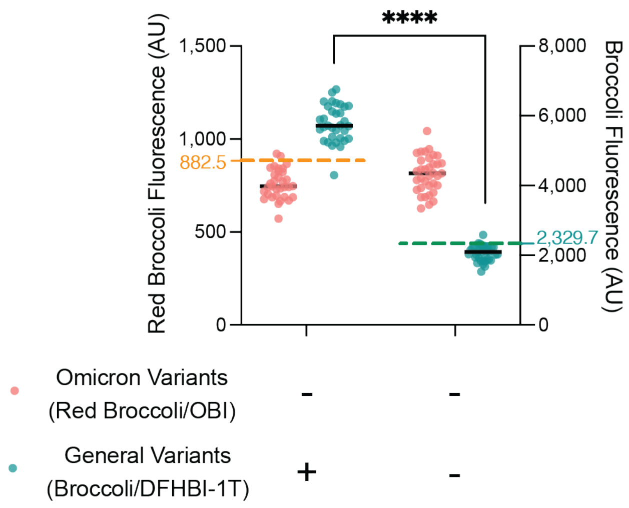

b

| | Negative by RT-qPCR | Positive b RT-qPCR |
|---|---|---|
| Positive by FARSIGHT | 0 | 12 |
| Negative by FARSIGHT | 11 | 0 |

| | Omicron by RNA-seq | Non Omicron by RNA-seq |
|---|---|---|
| Omicron by FARSIGHT | 12 | 0 |
| Non Omicron by FARSIGHT | 0 | 0 |

**Supplementary Fig. 12 l Control experiments and summary table for dual point-mutation detection assay by FARSIGHTs using clinical samples**

**a**, The 95th percentiles of value for Red Broccoli FARSIGHT non-template controls of Y505H(-)/N501Y(+) and Broccoli FARSIGHT Y505H(-)/N501Y(-) samples were used for threshold value determination (AU) of SARS-CoV-2 Omicron strains and general SARS-CoV-2 strains, respectively, at 2 hr.

**b**, Table summarizing performance of dual FARSIGHTs assay for recognizing the correct SARS-CoV-2 variants from 23 clinical patient saliva samples.

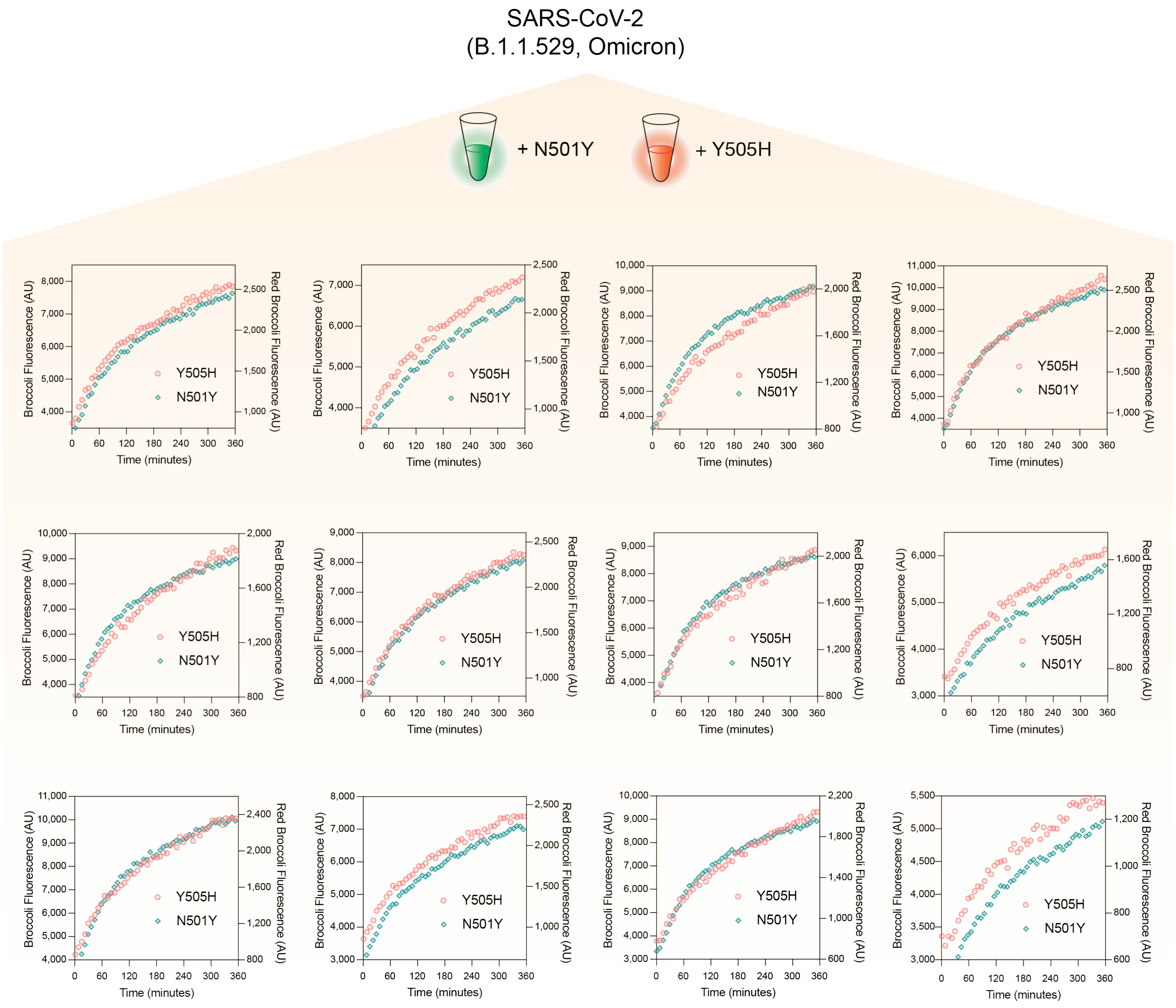

**Supplementary Fig. 13 I Time-course measurements of fluorescence from dual FARSIGHTs following NASBA with 12 positive Omicron BA.5 clinical saliva samples, Related to Fig. 6**

Red dots: fluorescence from Red Broccoli FARSIGHT targeting general SARS-CoV-2 Omicron variants (characterized by Y505H mutation).

Green dots: fluorescence from Broccoli FARSIGHT targeting general SARS-CoV-2 variants (characterized by N501Y mutation).

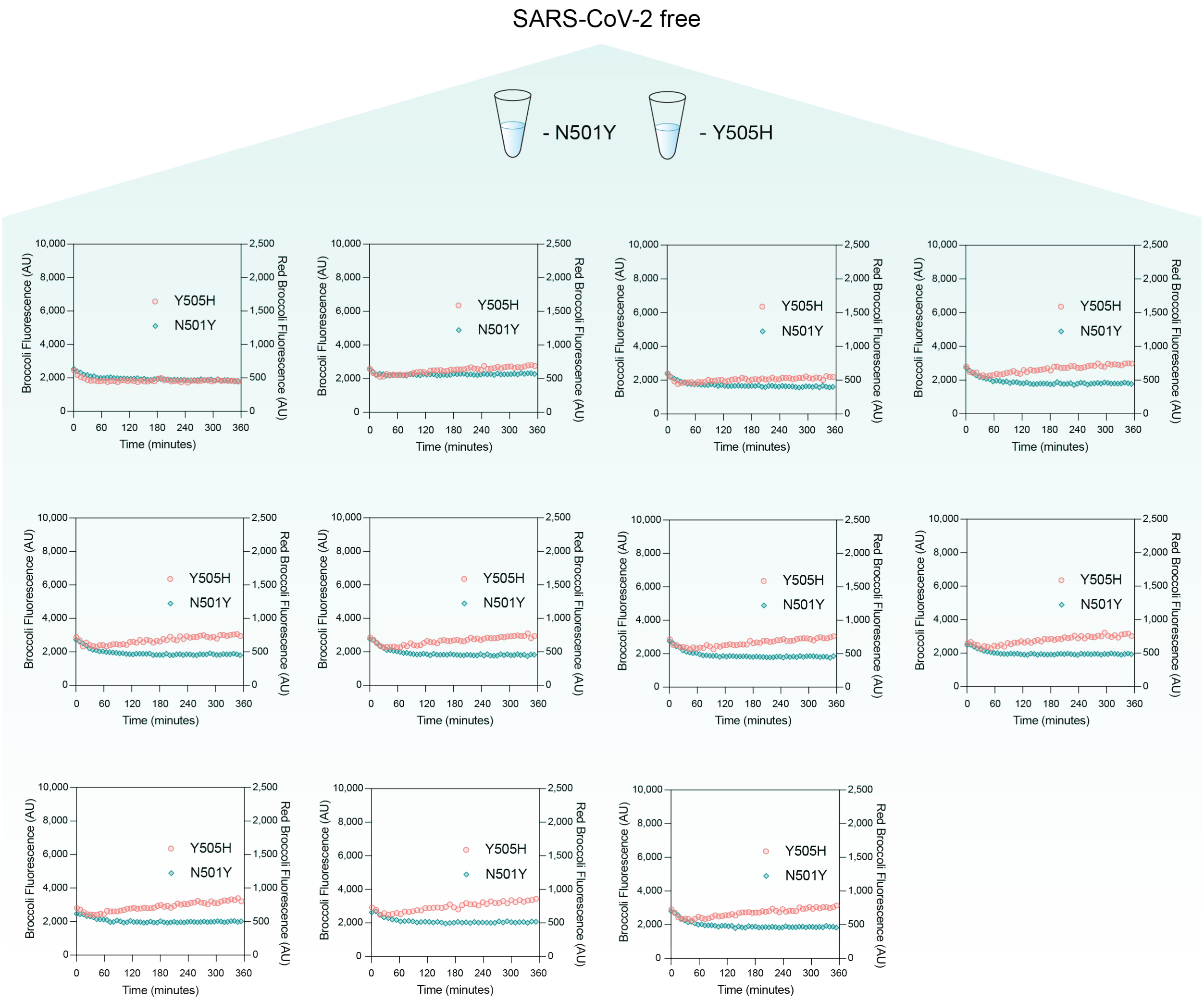

**Supplementary Fig. 14 | Fluorescence measurements of the dual FARSIGHT assay following NASBA with 11 negative clinical patient saliva samples, Related to Fig. 6**

Time-course measurements of fluorescence from dual FARSIGHTs following NASBA with 11 negative Omicron BA.5 clinical saliva samples.

Red dots: fluorescence from Red Broccoli FARSIGHT targeting general SARS-CoV-2 Omicron variants (characterized by Y505H mutation).

Green dots: fluorescence from Broccoli FARSIGHT targeting general SARS-CoV-2 variants (characterized by N501Y mutation).
